# Divergent treatment responses in chronic pain: Identifying subgroups of patients through cluster analysis

**DOI:** 10.1101/2024.02.23.24302234

**Authors:** Mienke Rijsdijk, Hidde M. Smits, Hazal R. Azizoglu, Sylvia Brugman, Yoeri van de Burgt, Tessa C. van Charldorp, Dewi J. van Gelder, Janny C. de Grauw, Eline A. van Lange, Frank J. Meye, Madelijn Strick, Hedi Walravens, Laura H.H. Winkens, Frank Huygen, Julia Drylewicz, Hanneke L.D.M. Willemen

## Abstract

**Background:** Chronic pain is an ill-defined disease with complex biopsychosocial aspects, posing treatment challenges. We hypothesize that treatment failure results, at least partly, from limited understanding of diverse patient subgroups. We aim to identify subgroups through psychometric data, allowing for more tailored interventions.

**Methods:** For this retrospective cohort study, we extracted patient-reported data from two Dutch tertiary multidisciplinary outpatient pain clinics (2018-2023) for unsupervised hierarchical clustering. Clusters were defined by anxiety, depression, pain catastrophizing, and kinesiophobia. Sociodemographics, pain characteristics, diagnosis, lifestyle, health-related quality of life (HRQoL) and treatment efficacy were compared among clusters. A prediction model was built utilizing a minimum set of questions to reliably assess cluster allocation.

**Results:** Among 5,454 patients with chronic pain, three clusters emerged. Cluster 1 (n=750) was characterized by high psychological burden, low HRQoL, lower educational levels and employment rates, and more smoking. Cluster 2 (n=1,795) showed low psychological burden, intermediate HRQoL, higher educational levels and employment rates, and more alcohol consumption. Cluster 3 (n=2,909) showed intermediate features. Pain reduction following treatment was least in cluster 1 (28.6% after capsaicin patch, 18.2% after multidisciplinary treatment), compared to >50% in clusters 2 and 3. A model incorporating 15 psychometric questions reliably predicted cluster allocation.

**In conclusion,** our study identifies distinct chronic pain patient clusters through 15 psychometric questions, revealing one cluster with notably poorer response to conventional treatment. Our prediction model may help clinicians improve treatment by allowing patient-subgroup targeted therapy according to cluster allocation.

**In brief:** Hierarchical clustering of chronic pain patients revealed three clusters based on pain experience and psychological welfare, with diverse sociodemographics and treatment effects suggesting potential for tailored interventions.

## Introduction

Every day, health care providers face challenges treating chronic pain patients, as treatment effects for this condition are often disappointing. The “numbers needed to treat (NNT)” for commonly used analgesic drugs, such as anti-neuropathic drugs and opioids, fall within the 3 to 10 range [1]. Recognizing that, depending on the drug, 3 to 10 patients need to be treated for a 50% pain reduction to be achieved in one patient, can be disheartening, as it entails treatment failure in the remaining patients. This is especially poignant as analgesic drugs prescribed for chronic pain can have serious side effects, such as opioid dependency and substance use disorder, which has contributed to the opioid crisis we are currently facing [2]. Hence, our approach to chronic pain treatment demands a transformation, and a potential solution involves deepening our understanding of the distinct characteristics found in clinical subgroups of patients experiencing chronic pain [3,4]. The identification of such subgroups could improve pain management by allowing treatment to be tailored to the needs and characteristics of each subgroup, ultimately reducing the NNT for specific analgesic interventions.

Identification of such patient subgroups should include biopsychosocial components, as chronic pain is a complex multi-faceted problem with important biological (e.g., genetics), psychological (e.g., anxiety, depression, pain catastrophizing), and social (e.g., low educational attainment, poor social support) factors all determining the experience of chronic pain. Several studies have supported the hypothesis that different subgroups of patients exist within different chronic pain populations, such as chronic low back pain [5], temporomandibular disorder [6] and fibromyalgia [7], whereas other studies have attempted to cluster patients in heterogenous chronic pain populations with a mix of painful conditions [8–12]. The available studies highly vary in their use of clustering variables, psychometric instruments and statistical methods. Clusters were based on unidimensional variables of pain-related characteristics such as pain location [9], or used a more multidimensional approach [8,10–12]. Combining the main results of these studies, 2 to 4 reliable clusters emerge with psychiatric symptoms such as anxiety and depression, as well as the psychological construct of catastrophizing, proving most important for cluster allocation. A clear relation with the biomedical domain (including pain diagnosis) and the social domain (including lifestyle, educational level and employment) is still missing. Likewise, it remains unclear whether different subgroups respond differently to some analgesic therapies than other subgroups.

The current study aimed to identify different chronic pain subgroups, incorporating aspects from all three dimensions of the biopsychosocial model. Data were derived from validated questionnaires that were used to assess mental and social health status variables in a heterogenous patient population in a tertiary outpatient pain clinic setting. The derived subgroups were compared in terms of sociodemographic characteristics, lifestyle factors, perceived health related quality of life (HRQoL), pain diagnosis, and treatment response. A secondary aim was to alleviate the burden on patients currently tasked with completing multiple (extensive) pain questionnaires, by identifying those questions that are essential for cluster allocation, and suggesting a concise questionnaire for this purpose.

## Methods

### Study design

In this retrospective observational cohort study all chronic pain patients referred to the tertiary multidisciplinary outpatient pain clinic of the University Medical Center Utrecht (UMCU) between May 2018 and May 2021, and of the Erasmus Medical Center Rotterdam (EMC), The Netherlands, between January 2017 and March 2023, were included. The Medical Research Ethics Committees of the UMCU (MEC-21/358) and of the EMC (MEC-2023-0161) both approved this study and waived the requirement to obtain informed consent.

### Data collection

Data were derived from questionnaires and standard entry boxes in the electronic health records that were collected as part of routine clinical care. Patients completed online questionnaires prior to their initial visit to the outpatient pain clinic. A combination of different patient-reported measures related to pain characteristics, psychological distress and health related quality of life (HRQoL) variables were included. These are described below.

#### Sociodemographics

Sociodemographic variables assessed were age (years), gender, Body Mass Index (BMI; kg/m²), lifestyle behaviors (alcohol consumption, drug use, smoking), having children, employment status, educational level, marital status and major life events (presence or absence, open to patients’ own definition).

#### Pain intensity, characteristics, duration and interference

*Pain intensity* was assessed using a 0 to 10 Numerical Rating Scale (NRS), with 0 equating to no pain and 10 to the worst imaginable pain, for the average, minimal and maximal pain intensity in the previous week.

We addressed *pain characteristics* in the UMCU using the first two questions of the Douleur Neuropathique en 4 (DN4) questionnaire comprising seven items (i.e., burning, painful cold, electric shock, pins and needles, tingling, numbness and pruritus) with a dichotomous yes-no scale. The total sum scores ranged from 0-7, with a cut-off point of ≥ 3 suggesting neuropathic pain [13]. The (Dutch) DN-4 has been validated in the general chronic pain population [14]. In the EMC, pain characteristics were assessed using the validated PainDetect, a 9-item self-report screening questionnaire [15]. It measures seven aspects of the quality of the pain experienced (i.e., burning, tingling, electric shocks, cold and heat hypersensitivity, numbness and pressure pain), the chronological pattern (time course), and whether or not the pain radiates. It is scored from 0 to 38, with total scores of less than 12 considered to represent nociceptive pain, 13–18 possible neuropathic pain, and scores ≥19 representing >90% likelihood of neuropathic pain.

For *pain duration*, patients indicated whether their pain persisted for more or less than one year.

*Pain interference* was assessed using the short form of Brief Pain Inventory (BPI) including seven items: general activity, mood, walking ability, normal work, relation with other people, sleep, and enjoyment of life. Each item was presented separately and was rated on a NRS scale from 0 to 10, with 0 indicating ‘no interference of pain with daily functioning’ and 10 ‘complete interference’[16,17].

#### Pain diagnosis

Patients were diagnosed during their first visit to the outpatient pain clinic by their attending anesthesiologist-pain specialist, shortly after filling out the questionnaires. Pain diagnoses were assessed according to the International Classification of Diseases 10 registry (ICD-10) [18].

#### Treatment effect

Treatment effect was assessed using the *Global Perceived Effect (GPE)* questionnaire. The GPE asks the patient to rate, on a 7-point Likert scale, how much their condition has improved or deteriorated since the start of treatment [19]. In the UMCU cohort a subgroup of patients with peripheral neuropathic pain or scar pain received a high concentration capsaicin 8% skin patch. Treatment effect was measured 14 days after capsaicin treatment. In the EMC cohort, treatment effect was measured three months after treatment initiation (multidisciplinary treatment) at the tertiary pain clinic.

#### Psychological distress variables

Psychological distress was measured using three different questionnaires. The *Hospital Anxiety and Depression Scale (HADS)* self-assessment questionnaire assesses the level of anxiety and depression symptoms [20,21]. The *Pain Catastrophizing Scale (PCS)* assesses catastrophizing in three dimensions: magnification, rumination and helplessness [22–24]. The *Tampa Scale of Kinesiophobia (TSK)* questionnaire assesses fear of movement and injury [25].

#### Health-related Quality of life

In the UMCU cohort, patients completed either the European Quality of Life instrument (EQ5D) or 12-item Short Form Health Survey (SF-12), because clinical practice changed during the study period, with a switch from the EQ5D to the SF-12. In the EMC cohort, the 36-item Short Form Health Survey (SF-36) was used. Details of the abovementioned questionnaires are reported in the Supplementary file to this methods section.

### Statistical analyses

A statistical analysis plan was formalized before accessing the data for the primary outcome. No statistical power calculation was conducted prior to the study and all available data were included. Hierarchical clustering was performed on psychometric data using the individual questions of the HADS-A, HADS-D, PCS and TSK questionnaires. We chose these questionnaires as they were the only questionnaires used by both study centres. We decided to leave HRQoL out of the cluster analysis, as patients filled out either the EQ5D, the SF12, or the SF-36, which would lead to exclusion of a large number of patients.

All patients with at least one missing value for one of the questions were excluded from this analysis. No imputation of missing data was performed as this could influence the clustering analysis.

Cluster analysis was performed using squared Euclidean distances to determine the similarity and Ward’s minimum variance as the clustering method to minimize within-cluster differences.

The derived clusters were compared for pain intensity, duration, characteristics and interference, pain diagnosis, sociodemographic variables, and HRQoL using one-way ANOVA for normally distributed continuous variables and Pearsons-Chi-square tests for categorical variables. Continuous data were expressed as mean with 95% confidence intervals, categorical data as counts and percentages, and medians with interquartile range were chosen for NRS data. Statistical significance was set at p ≤ 0.01 to account for multiple testing. Subsequently, effect sizes of the observed significant differences were estimated using eta squared with <0.06 classified as small, 0.06 to 0.14 as medium and ≥ 0.14 as a larger effect size, or using Cramér’s V with 0.1 to 0.3 as a small, 0.3 to 0.5 as a medium, and ≥ 0.5 as a large effect size (47).

The random forest model was used as a prediction model for which the UMCU cohort was the discovery cohort and the EMC cohort acted as the validation cohort.

Differences in treatment effect were based on the results of the GPE and percentage of change in the NRS score between baseline and follow-up. Answers to the GPE were reduced to a dichotomous variable of “improved” (including little improvement, much improvement, and fully recovered) or “not improved” (including unchanged, little worse, much worse, and very bad) after treatment. Differences in treatment effects were analysed using a Pearsons-Chi-square test with improvement yes/no as outcome parameter, and a paired T-test for pain decrease (NRS) as outcome measure. Statistical analyses were performed using IBM SPSS Statistics version 26.0 to compare derived clusters for pain intensity, duration, characteristics and interference, pain diagnosis, sociodemographic variables, and HRQoL) and R version 4.2.2.

## Results

### Sample description

In total, 8,133 patients were included in the study, of whom 2,654 were referred to the UMCU and 5,479 to the EMC. Of these, 1,043 (UMCU; 39%) and 1,636 (EMC; 30%) were excluded due to one or more missing values in the questionnaires used for cluster analysis, resulting in 1,611 UMCU patients and 3,843 EMC patients in the final analysis (Fig 1).

**Fig 1.**
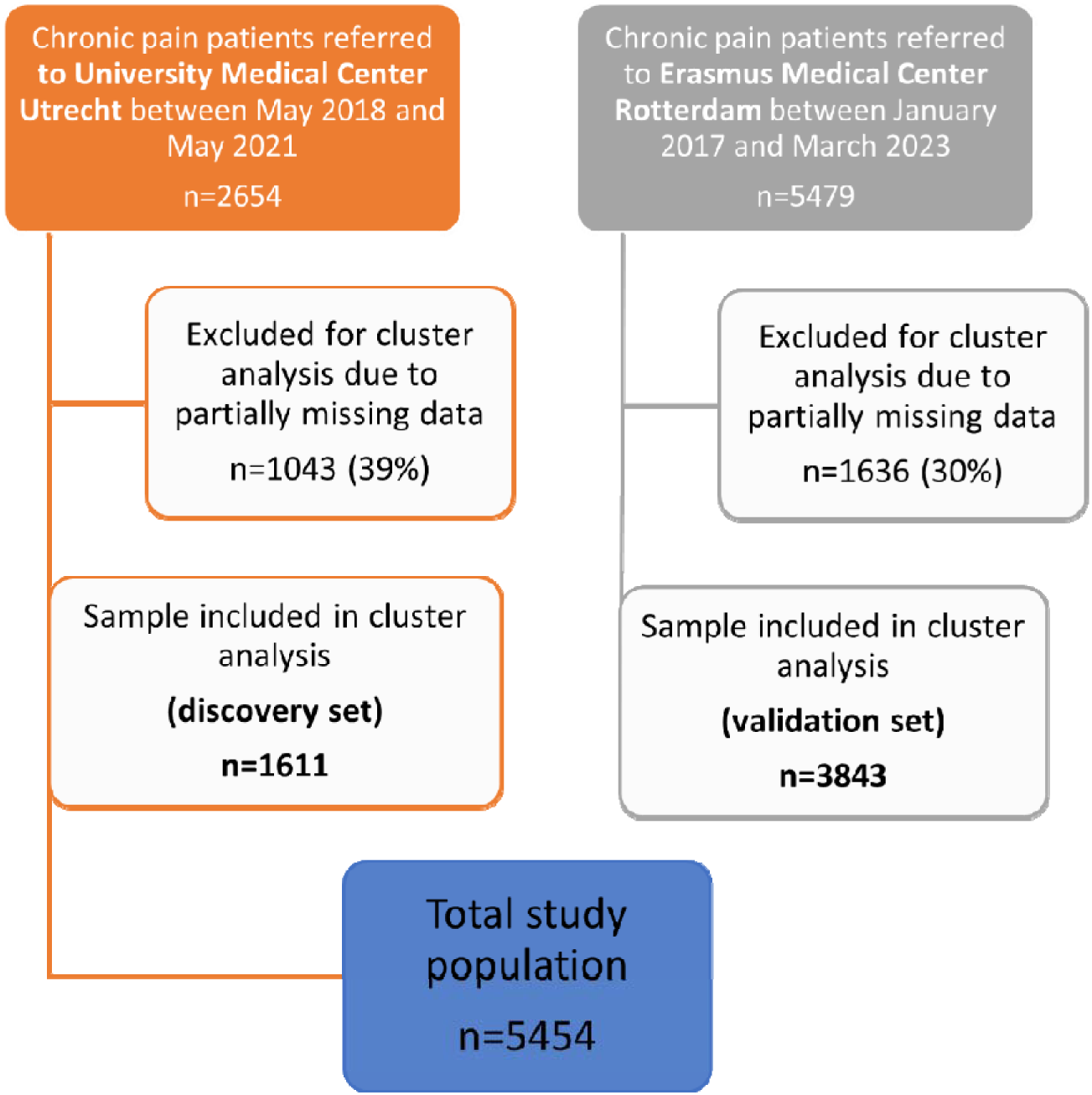
Flowchart of study population.

The patients in the UMCU cohort were slightly older than in the EMC cohort (mean age 54.5 years (95% CI 53.7-55.4) versus 49.9 (95% CI 49.4 -50.4)). There were fewer females included in the UMCU population (53.9% females versus 64.2% in the EMC). The majority in both cohorts was married or cohabiting (UMCU 77.3% and EMC 75.0%) and had children (UMCU 71.5% and EMC 68.2%). A minority of patients was employed (UMCU 35.8% and EMC 37.1%) (Table 1).

**Table 1.**
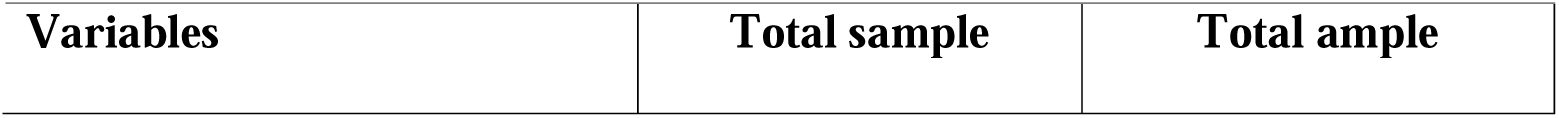
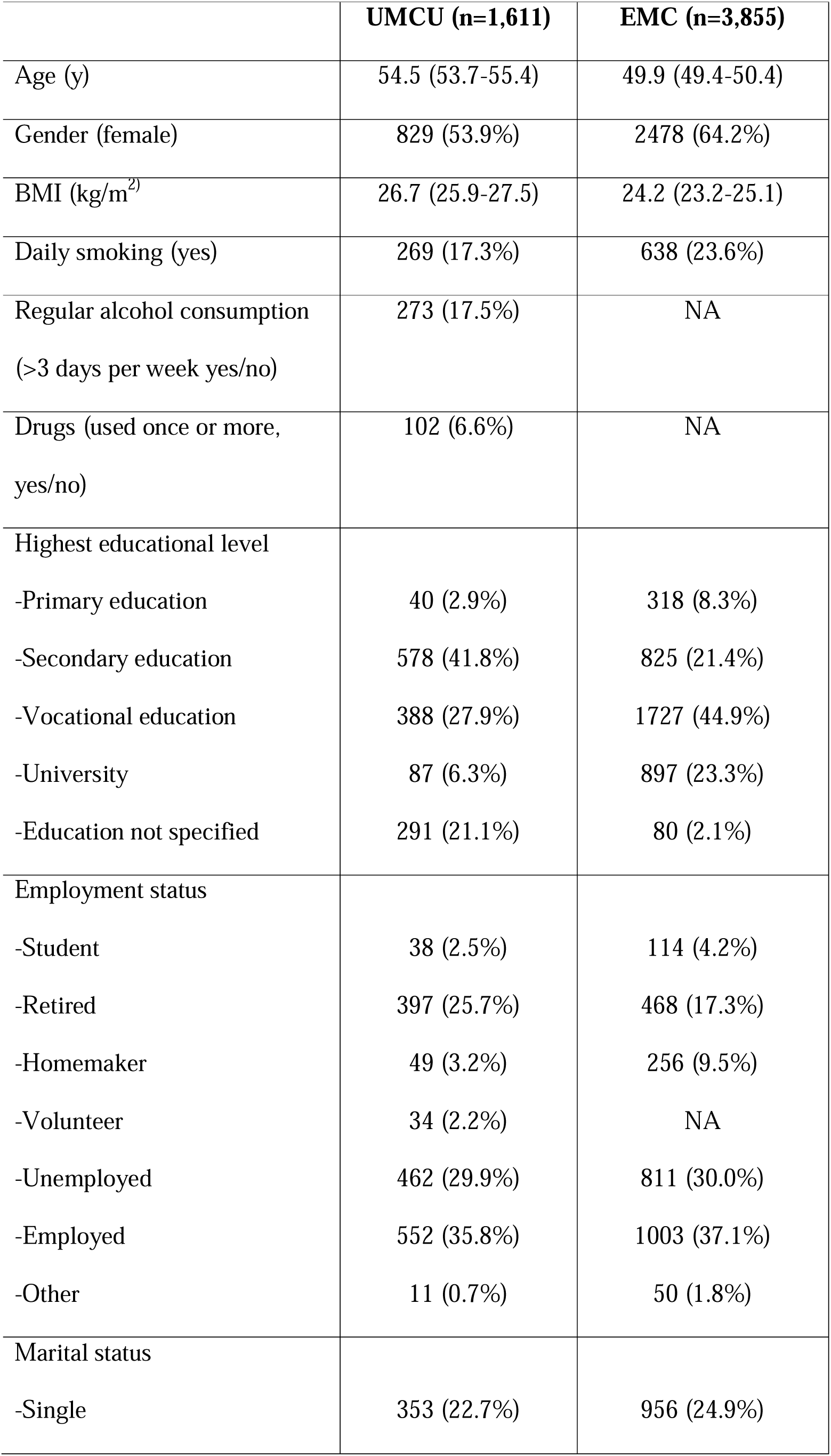
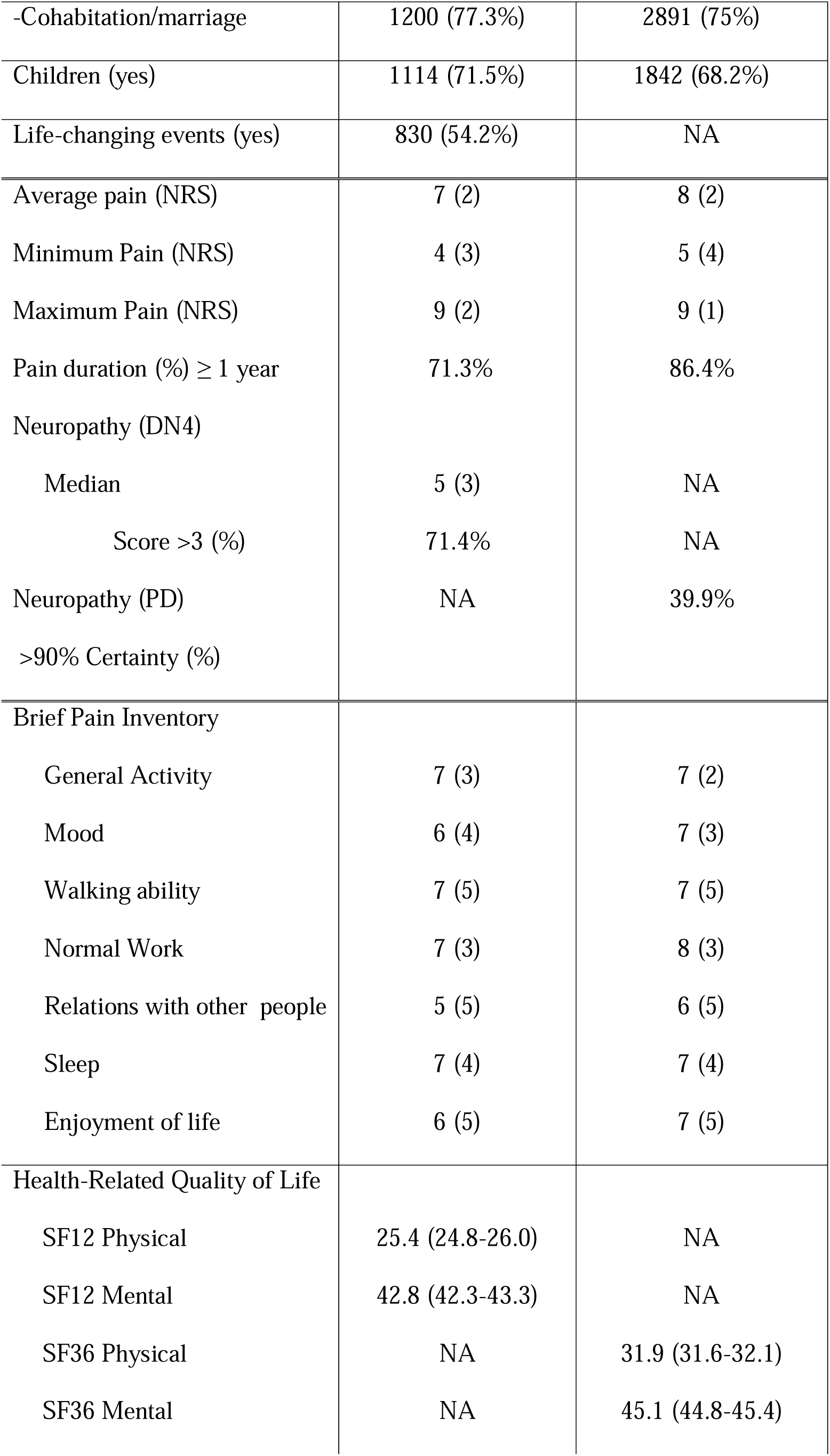
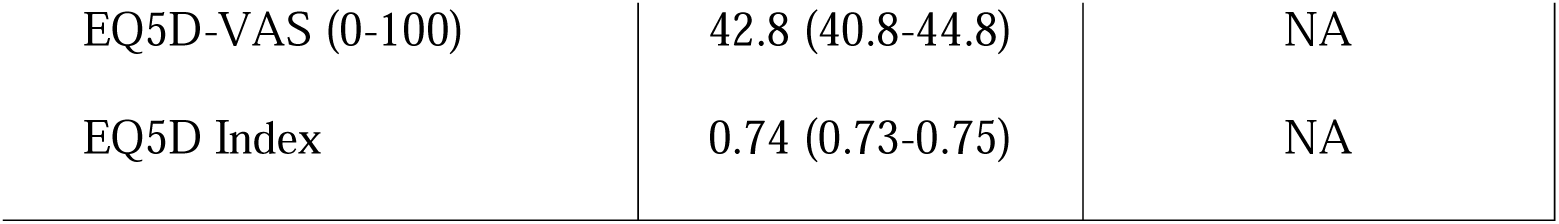
Sociodemographics, pain intensity, duration, character, interference and health-related quality of life in the study population (University Medical Center Utrecht and Erasmus Medical Center Rotterdam). Data expressed as mean (95% confidence interval), median (IQR) or count (%). BMI: Body Mass Index; DN4: Douleur Neuropathique en 4; EMC: Erasmus Medical Center Rotterdam; EQ5D: European Quality of Life instrument 5; EQ5DVAS: European Quality of Life instrument 5-Visual Analogue Scale; NRS: Numeric Rating Scale; PD: PainDetect; SF12: Short Form-12; SF36: Short Form-36; UMCU: University Medical Center Utrecht.

|  | **UMCU (n=1,611)** | **EMC (n=3,855)** |
| --- | --- | --- |
| Age (y) | 54.5 (53.7-55.4) | 49.9 (49.4-50.4) |
| Gender (female) | 829 (53.9%) | 2478 (64.2%) |
| BMI (kg/m<sup>2</sup>) | 26.7 (25.9-27.5) | 24.2 (23.2-25.1) |
| Daily smoking (yes) | 269 (17.3%) | 638 (23.6%) |
| Regular alcohol consumption<br>(>3 days per week yes/no) | 273 (17.5%) | NA |
| Drugs (used once or more,<br>yes/no) | 102 (6.6%) | NA |
| Highest educational level |  |  |
| -Primary education | 40 (2.9%) | 318 (8.3%) |
| -Secondary education | 578 (41.8%) | 825 (21.4%) |
| -Vocational education | 388 (27.9%) | 1727 (44.9%) |
| -University | 87 (6.3%) | 897 (23.3%) |
| -Education not specified | 291 (21.1%) | 80 (2.1%) |
| Employment status |  |  |
| -Student | 38 (2.5%) | 114 (4.2%) |
| -Retired | 397 (25.7%) | 468 (17.3%) |
| -Homemaker | 49 (3.2%) | 256 (9.5%) |
| -Volunteer | 34 (2.2%) | NA |
| -Unemployed | 462 (29.9%) | 811 (30.0%) |
| -Employed | 552 (35.8%) | 1003 (37.1%) |
| -Other | 11 (0.7%) | 50 (1.8%) |
| Marital status |  |  |
| -Single | 353 (22.7%) | 956 (24.9%) |
| -Cohabitation/marriage | 1200 (77.3%) | 2891 (75%) |
| Children (yes) | 1114 (71.5%) | 1842 (68.2%) |
| Life-changing events (yes) | 830 (54.2%) | NA |
| Average pain (NRS) | 7 (2) | 8 (2) |
| Minimum Pain (NRS) | 4 (3) | 5 (4) |
| Maximum Pain (NRS) | 9 (2) | 9 (1) |
| Pain duration (%) ≥ 1 year | 71.3% | 86.4% |
| Neuropathy (DN4) |  |  |
| Median | 5 (3) | NA |
| Score >3 (%) | 71.4% | NA |
| Neuropathy (PD) | NA | 39.9% |
| >90% Certainty (%) |  |  |
| Brief Pain Inventory |  |  |
| General Activity | 7 (3) | 7 (2) |
| Mood | 6 (4) | 7 (3) |
| Walking ability | 7 (5) | 7 (5) |
| Normal Work | 7 (3) | 8 (3) |
| Relations with other people | 5 (5) | 6 (5) |
| Sleep | 7 (4) | 7 (4) |
| Enjoyment of life | 6 (5) | 7 (5) |
| Health-Related Quality of Life |  |  |
| SF12 Physical | 25.4 (24.8-26.0) | NA |
| SF12 Mental | 42.8 (42.3-43.3) | NA |
| SF36 Physical | NA | 31.9 (31.6-32.1) |
| SF36 Mental | NA | 45.1 (44.8-45.4) |
| EQ5D-VAS (0-100) | 42.8 (40.8-44.8) | NA |
| EQ5D Index | 0.74 (0.73-0.75) | NA |

In the EMC cohort, more patients had experienced pain for more than 1 year (86.4% versus 71.3% in the UMCU cohort), with no difference in pain intensity between the cohorts (median NRS in the past week in UMCU cohort was 7 (IQR= 5-9) versus 8 (IQR= 6-10) in the EMC). We cannot meaningfully compare presence of neuropathic pain characteristics between both cohorts as two different questionnaires (DN4 and PainDetect) were used (Table 1). In the UMCU cohort, the most common diagnoses were radicular syndrome (24.7%), mechanical spine related pain (11.6%), and mononeuropathy (8.1%) (Table 2). In the EMC, this was “other neuropathic pain“ (17.5%), radicular syndrome (17.0%) and tendomyogenic pain (12.1%) (S1 Table).

**Table 2.**
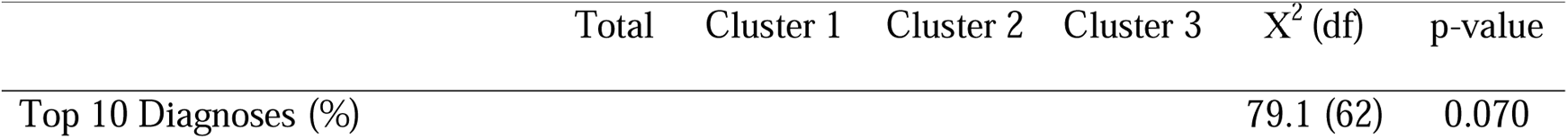
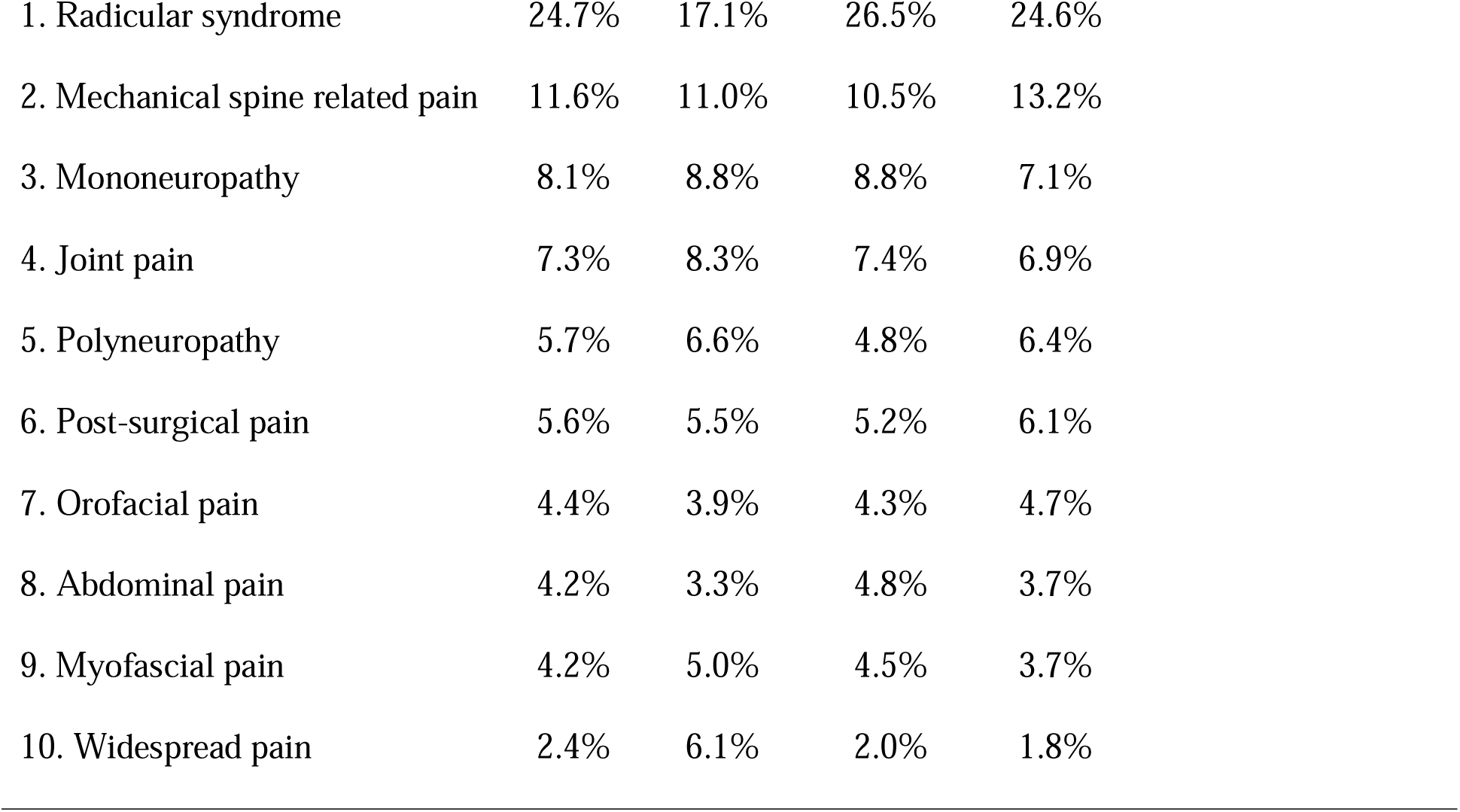
Legend. Frequencies of the 10 most common diagnoses in each cluster for the <u>discovery cohort (UMCU).</u>. Data expressed as percentage per cluster. Significance levels computed by Pearson-Chi square. There is no missing data.

|  | Total | Cluster 1 | Cluster 2 | Cluster 3 | X<sup>2</sup> (df) | p-value |
| --- | --- | --- | --- | --- | --- | --- |
| Top 10 Diagnoses (%) |  |  |  |  | 79.1 (62) | 0.070 |

|  |  |  |  |  |
| --- | --- | --- | --- | --- |
| 1. Radicular syndrome | 24.7% | 17.1% | 26.5% | 24.6% |
| 2. Mechanical spine related pain | 11.6% | 11.0% | 10.5% | 13.2% |
| 3. Mononeuropathy | 8.1% | 8.8% | 8.8% | 7.1% |
| 4. Joint pain | 7.3% | 8.3% | 7.4% | 6.9% |
| 5. Polyneuropathy | 5.7% | 6.6% | 4.8% | 6.4% |
| 6. Post-surgical pain | 5.6% | 5.5% | 5.2% | 6.1% |
| 7. Orofacial pain | 4.4% | 3.9% | 4.3% | 4.7% |
| 8. Abdominal pain | 4.2% | 3.3% | 4.8% | 3.7% |
| 9. Myofascial pain | 4.2% | 5.0% | 4.5% | 3.7% |
| 10. Widespread pain | 2.4% | 6.1% | 2.0% | 1.8% |

### Hierarchical clustering revealed 3 clusters of patients

We first performed a hierarchical clustering on the combined datasets (UMCU and EMC) using the individual questions of the HADS-A, HADS-D, PCS and TSK questionnaires. We identified 3 distinct clusters of patients: Cluster 1 included 750 patients (13.8%), Cluster 2 1,795 patients (32.9%), and Cluster 3 comprised of 2,909 patients (53.3%) (Fig 2). When performing separate hierarchical clustering on the datasets (i.e., UMCU data separate from EMC data), we found that more than 75% of the patients were assigned to the same cluster (not shown).

**Fig 2.**
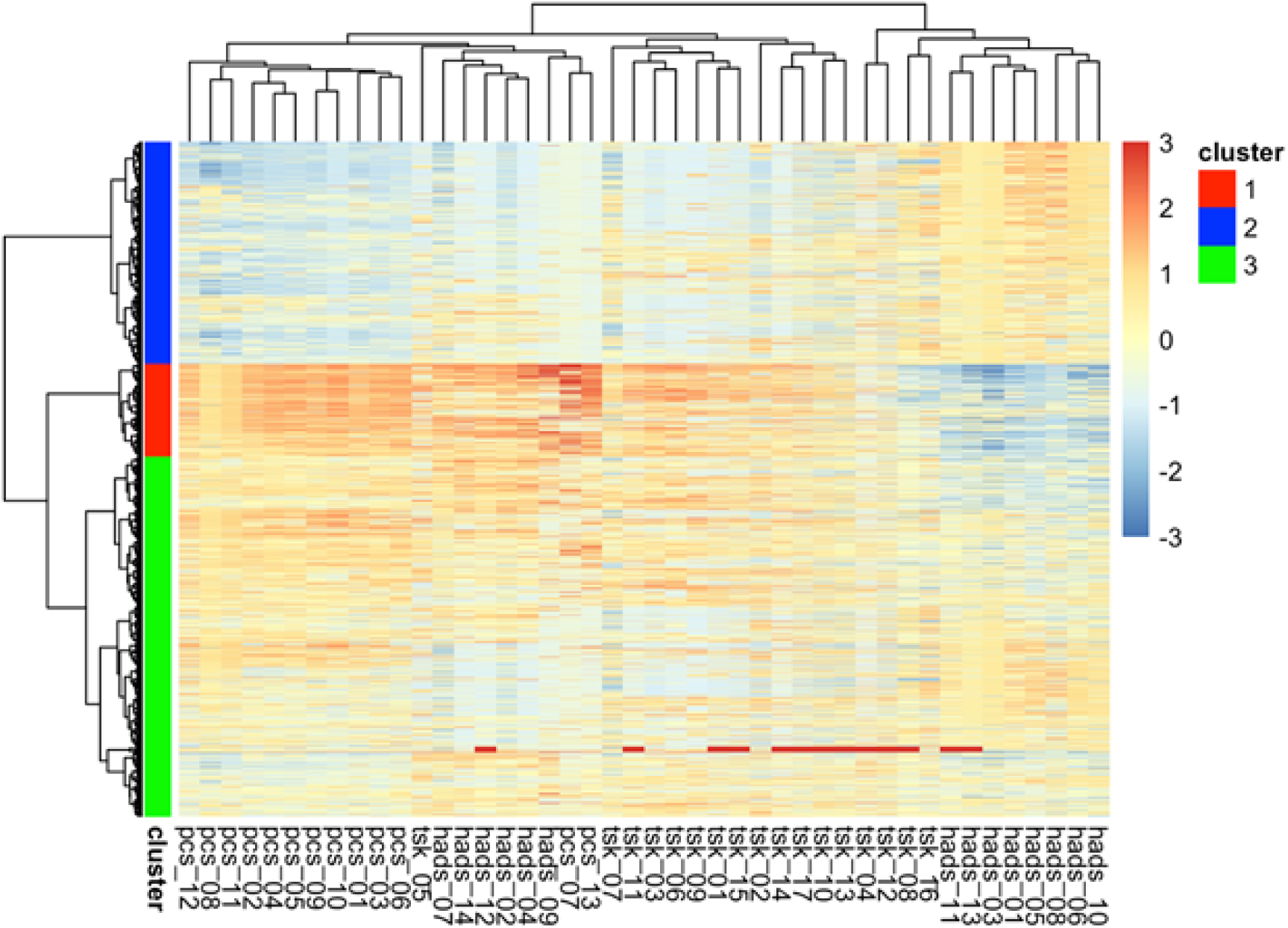
Hierarchical clustering dendogram of the cohorts combined (UMCU and EMC). Hierarchical cluster analysis showing three clusters: Cluster 1(red), Cluster 2 (blue) and Cluster 3 (green). X-axis representing the individual questions of three questionnaires (Hospital Anxiety and Depression Scale (HADS), Pain Catastrophizing Scale (PCS), and Tampa Scale of Kinesiophobia (TSK). Y-axis representing included patients.

Cluster 1 was characterized by higher scores for anxiety (HADS-A mean 13.4; 95%CI=13.1-13.7), depression (HADS-D mean 13.6 (95%CI=13.3-13.9)), catastrophizing (PCS mean 43.6; 95%CI=43.2-44.0) and kinesiophobia or pain related fear (TSK mean 46.9; 95%CI=46.4-47.4). Cluster 2 on the other hand was characterized by the lowest scores for each of these characteristics (HADS-A mean 3.7; 95%CI=3.6-3.8, HADS-D mean 4.1; 95%CI=4.0-4.3, PCS mean 10.3; 95%CI=10.0-10.6 and TSK 34.0; 95%CI=33.7-34.3), while cluster 3 showed intermediate scores, at or just passing the cut-off scores for anxiety, depression, catastrophizing and kinesiophobia (HADS-A 7.1; 95%CI=6.9-7.2, HADS-D 7.9; 95%CI=7.7-8.0, PCS 26.8; 95%CI=26.5-27.1 and TSK 40.1; 95%CI=39.9-40.4) (Table 3).

**Table 3.** Clustering characteristics in the total study population (UMCU and EMC) and comparison among the clusters. Data expressed as mean (95% confidence interval) or percentage (%) of count data. Statistics computed by one-way ANOVA and effect sizes are calculated with eta squared with <0.06 as small, 0.06 to 0.14 as medium and ≥0.14 as a larger effect size, EMC: Erasmus Medical Center Rotterdam; HADS-A and HADS-D: Hospital Anxiety and Depression Scale; PCS: Pain Catastrophizing Scale; TSK: Tampa Scale for Kinesiophobia; UMCU: University Medical Center Utrecht. PCS subcategories represent the sum of following items ‘Rumination’ 8,9, 10 and 11; Magnification 6, 7 and 13; ‘Helplessness’ 1, 2, 3, 4, 5 and 12. Percentage of missing data: HADS 0%, PCA 0%, TSK 0%

| Variables | Total sample<br>(n=5454) | Cluster 1<br>(n=750, 13.8%) | Cluster 2<br>(n=1795, 32.9%) | Cluster 3<br>(n=2909, 53.3 %) | *p*-value | *Effect size*<br>*Eta-squared* |
| --- | --- | --- | --- | --- | --- | --- |
| Anxiety (HADS-A) | 6.8 (6.7-7.0) | 13.4 (13.1-13.7) | 3.7 (3.6-3.8) | 7.1 (6.9-7.2) | <0.001 | **0.455** |
| -Signs of anxiety disorder (Score >8 (%)) | 30.6% | 87.3% | 3.7% | 32.7% |  |  |
| -Clinical significant anxiety disorder (Score >10 (%)) | 20.4% | 78.3% | 1.0% | 17.4% |  |  |
| Depression (HADS-D) | 7.4 (7.3-7.6) | 13.6 (13.3-13.9) | 4.1 (4.0-4.3) | 7.9 (7.7-8.0) | <0.001 | **0.373** |
| -Signs of depression (Score >8 (%)) | 39.3% | 89.5% | 11.8% | 43.4% |  |  |
| -Clinical significant depression (Score >10 (%)) | 27.4% | 78.3% | 5.0% | 28.2% |  |  |
| Pain catastrophizing<br>(PCS) Total score | 23.7 (23.3-24.0) | 43.6 (43.2-44.0) | 10.3 (10.0-10.6) | 26.8 (26.5-27.1) | **<0.001** | **0.703** |
| Rumination | 8.8 (8.7-9.0) | 14.5 (14.4-14.6) | 4.5 (4.3-4.6) | 10.1 (10.0-10.2) |  |  |
| Magnification | 3.4 (3.3-3.4) | 8.5 (8.3-8.7) | 0.9 (0.8-0.9) | 3.6 (3.5-3.7) |  |  |
| Helplessness | 11.5 (11.3-11.6) | 20.6 (20.4-20.8) | 5.0 (4.8-5.1) | 13.1 (13.0-13.3) |  |  |
| Pain-related fear (TSK) | 39.0 (38.8-39.2) | 46.9 (46.4-47.4) | 34.0 (33.7-34.3) | 40.1 (39.9-40.4) | **<0.001** | **0.273** |

Signs of a clinically significant anxiety and/or depression disorder (HADS-A and HADS-D above 12, for which patients may be referred to a psychiatrist for additional diagnostic testing) were far more frequently observed in cluster 1 (78.3% anxiety disorder and 78.3% depression disorder; p-value <0.001) compared to cluster 2 (anxiety disorder 1.0% and depression disorder 5%, p-value <0.001) and cluster 3 (anxiety disorder 17.4% and depression disorder 28.2%; p-value <0.001) (Table 3).

### Differences between the clusters in sociodemographic characteristics, lifestyle behaviors, and pain characteristics

Regarding *sociodemographic characteristics and lifestyle behaviors*, patients in cluster 1 smoked tobacco more often, and were more often single. Patients in cluster 2 consumed more alcohol (in UMCU cohort; this was not recorded in the EMC cohort), had the highest educational levels, and the highest employment rates (Table 4, and S2 Table).

**Table 4.**
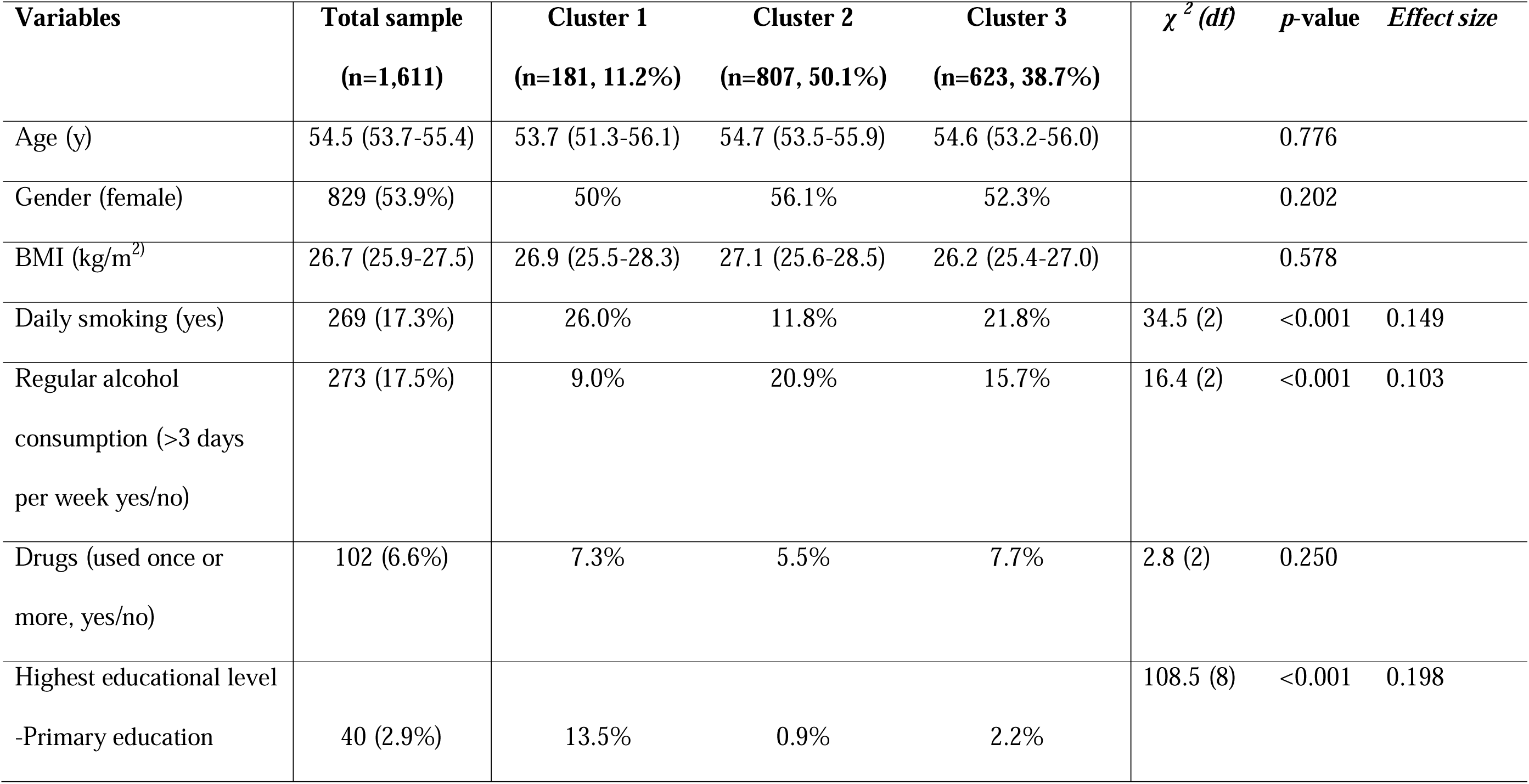
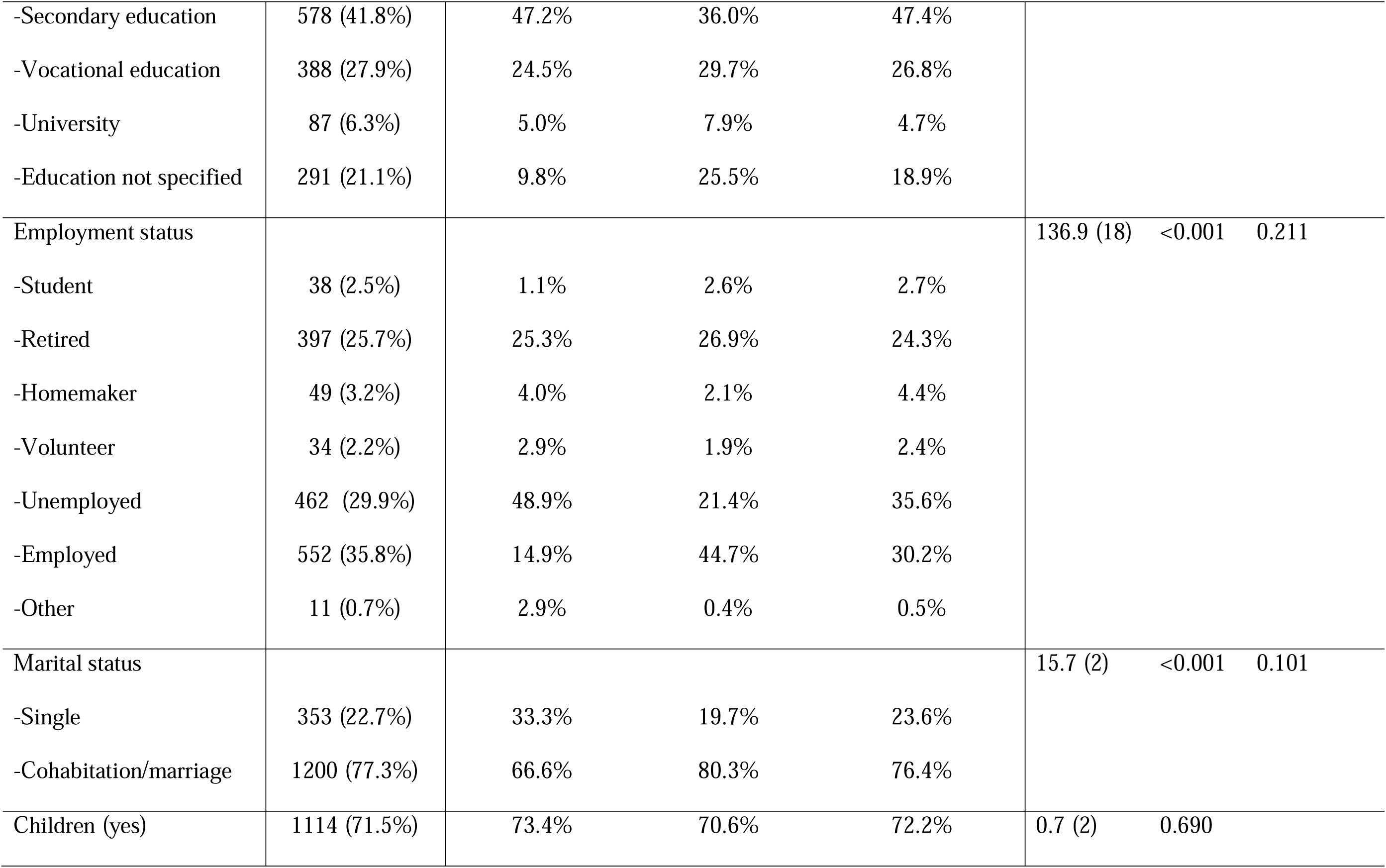
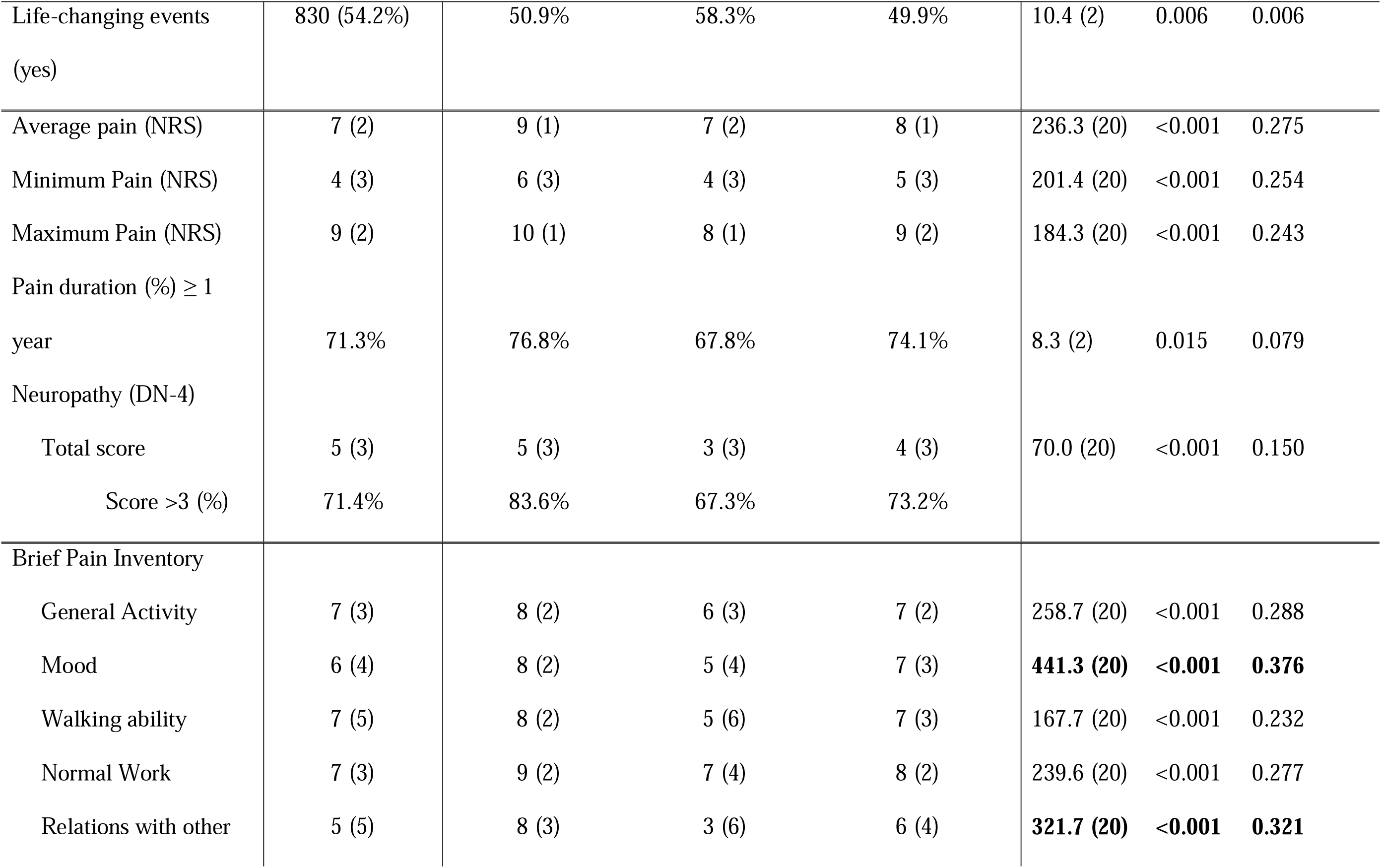
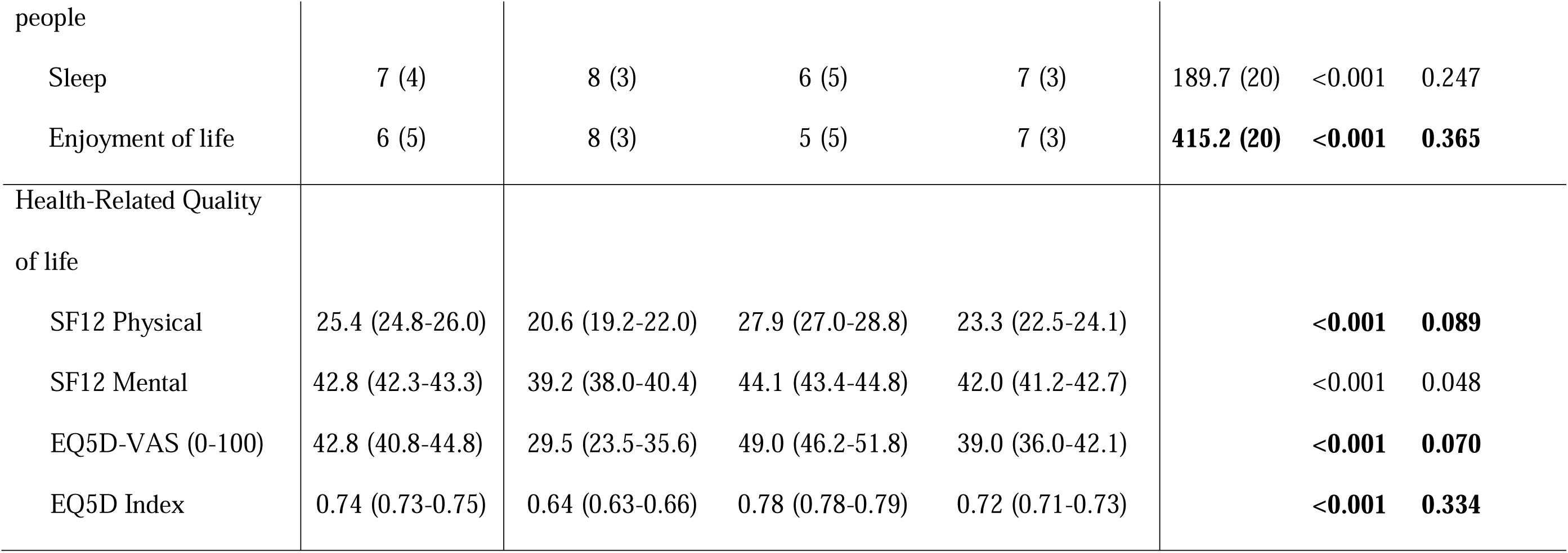
Sociodemographics, pain intensity, duration, character, interference and health-related quality of life in the discovery cohort (UMCU) and comparison among the clusters. Sociodemographic data expressed as mean (95% confidence interval) or count (%). Questionnaire scores are expressed as median (interquartile range) or count (%). Statistics computed by one-Way ANOVA Test or Pearson Chi-Square test. Effect sizes for continuous data were estimated using eta squared with <0.06 as small, 0.06 to 0.14 as medium and ≥0.14 as a larger effect size. Effect sizes for categorical data are calculated with Cramer’s V. Significance levels computed by Pearson Chi-Square test and effect sizes with Cramer’s V indicating small 0.1 to 0.3, medium 0.3 to 0.5 and large >0.5 effect sizes. **Significant differences with medium to large effect size are in bold.** Percentage of missing data: age 0.3%, gender 4.6%, BMI 76.0%, daily smoking 3.4%, regular alcohol 3.4%, drugs 3.4%, highest educational level 14.3%, employment status 4.3%, marital status 3.7%, children 3.4%, life-changing events 5.1%, pain NRS 0.2%, DN4 0,2%, BPI items 0.2%, TSK, HADS 0.3%, PCS 0.2%, SF12 43.8%, EQ5D 59.5%. BMI: Body Mass Index; DN4: Douleur Neuropathique en 4; EQ5D: European Quality of Life instrument 5; EQ5D’-VAS: European Quality of Life instrument 5-Visual Analogue Scale; NRS: Numeric Rating Scale; SF12: Short Form-12; UMCU: University Medical Center Utrecht.

In the UMCU cohort, *pain* was most severe in cluster 1 with the highest intensity (median pain during the past week NRS 9; IQR= 8-10), longest duration (76.8% over one year) and highest prevalence of neuropath (83.6%). Cluster 2 showed lowest pain severity scores (NRS 7; IQR= 5-9, pain duration > 1 year in 67.8% of patients, 67.3% show signs of neuropathy) and cluster 3 intermediate scores (NRS 8; IQR= 7-9, pain duration > 1 year in 74.1% of patients, 73.2% signs of neuropathy). Differences were significant between groups (Table 4).

In the EMC cohort, some different observations were made. Pain intensity and duration were comparable between clusters. Signs of neuropathic pain however were most often observed in cluster 2 (50.8%), compared with31.9% in cluster 1 and 39.5% in cluster 3. All differences had a small effect size (S2 Table).

In cluster 1, widespread pain (UMCU cohort) and tendomyogenic pain (EMC cohort) were most prevalent, and radicular syndrome least prevalent (both cohorts). Complex regional pain syndrome (EMC cohort) was most prevalent in cluster 2. For all other diagnoses, there were no clinically relevant differences in prevalence among clusters (Table 2, S1 Table). Pain influenced HRQoL most in cluster 1 in both cohorts (Table 4, S2 Table).

### Treatment efficacy differs between clusters

Next, we tested whether medical treatment efficacy was different between the three subgroups. In n=104 patients referred to the UMCU and receiving a Capsaicin 8% patch for peripheral neuropathic pain or scar pain, 28.6% of those in cluster 1 experienced improvement at 14 days follow up after treatment, compared to, 58.9% and 55.9% in cluster 2 and 3 respectively. In cluster 1, although patients reported a small improvement on the GPE, maximum pain scores did not decrease significantly (NRS mean 8.9 (95%CI=8.4-9.5) to 8.6 (95%CI=8.0-9.3), p=0.391) at 14 days follow-up, while in cluster 2 and 3 a significant reduction in maximum pain scores was observed (cluster 2 NRS mean 8.1 (95%CI=7.8-8.4) to 6.2 (95%CI=5.5-6.9), p<0.001; cluster 3 NRS mean 8.2 (95%CI=7.8-8.5) to 6.5 (95%CI=5.7-7.3), p<0.001) (Table 5).

**Table 5.**
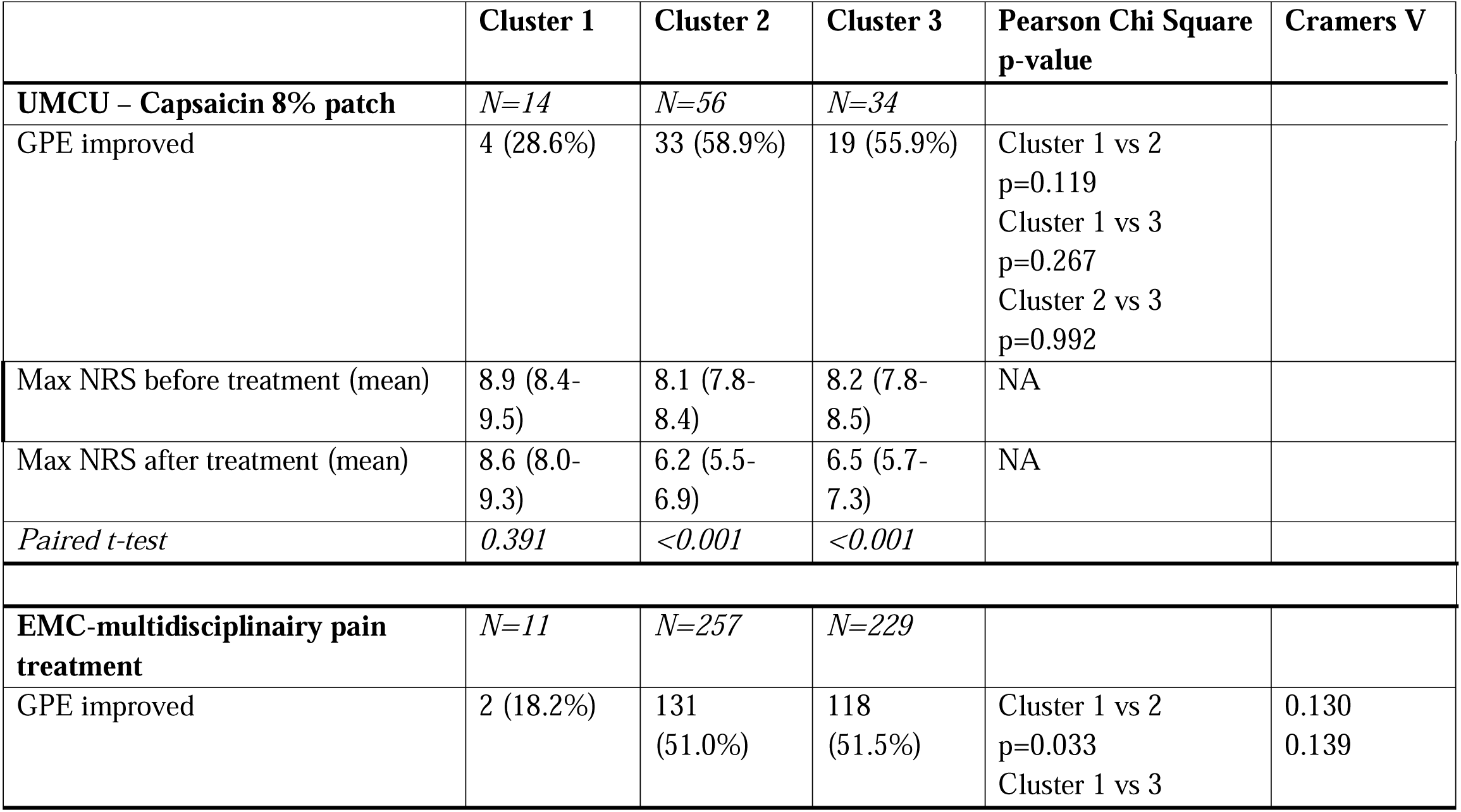
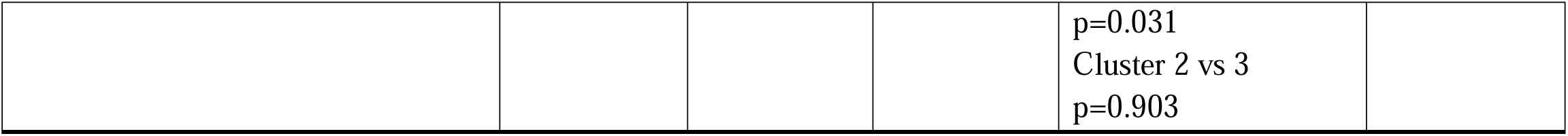
Treatment effect differs between cluster. Treatment effect (global perceived effect and pain numeric rating scale) were assessed two weeks after application of a capsaicin 8% patch in patients with peripheral neuropathic pain or scar pain in a subgroup of patient in the UMCU cohort. Treatment effect (global perceived effect) was assessed three months after baseline after a multidisciplinary treatment in the EMC cohort. All available data was used. The global perceived effect was categorized to “improved” of “not improved (including no change)”. Data expressed as count (%).Significance levels computed by Pearson Chi-Square test and effect sizes with Cramer’s V indicating small 0.1 to 0.3, medium 0.3 to 0.5 and large >0.5 effect sizes. The pain numeric rating scale was calculated as mean and baseline and follow up NRS scores were calculated within clusters using a paired-sample t-test. P-values <0.05 were regarded as significant.

Treatment outcome was assessed three months after baseline in n=497 (12.9%) patients who were referred to the EMC and received multidisciplinary pain treatment (all treatment options in the EMC are included). Improvement was reported in 18.2% of patients in cluster 1, compared to 51.0% in cluster 2, and 51.5% in cluster 3 (Table 5).

### Cluster membership prediction is accurate using only 15 questions

In a post-hoc analysis, we next aimed to predict cluster membership by using the UMCU dataset as a discovery cohort and the EMC dataset as a validation cohort. Each patient (from the discovery and validation cohort) was assigned to the cluster defined in the analysis pooling both datasets together. We used a random forest approach to classify the patients of the discovery cohort and reached an overall accuracy of 86%, with overall high sensitivity and specificity for each cluster (S3A Table). The defined prediction model was then used on the validation cohort and the predicted memberships were compared to the ones assigned by the previously performed hierarchical clustering. When using all the questions (n=44), the accuracy was 72.6% and cluster specific sensitivity and specificity were high for cluster 1 and 2 (S3A Table).

To reduce patients’ burden in having to answer multiple lengthy questionnaires, we next investigated whether we could reduce the number of questions and still obtain adequate accuracy, sensitivity and specificity for the prediction model. An advantage of a random forest model is that it tells one which variables are most important to accurately predict classes. Hence, we again used the validation cohort, but this time with only the 20 most important questions (S1 Fig). This new model showed an accuracy of 70.3%, again with good sensitivity and specificity (S3B Table). Using the 15 most important questions resulted in an accuracy of 70.0% (S3C Table), while using the 10 most important questions yielded an accuracy of 69.6% but lower balanced accuracy (S3D Table). Overall, using the 15 most important questions seems to provide the best balance between number of questions and desired prediction accuracy. The balanced accuracy with 15 questions for clusters 1, 2 and 3 was 91%, 84% and 68% respectively. The prediction model has an approximate sensitivity for cluster 1, 2 and 3 of 96%, 74% and 58%, and specificity of 86%, 95% and 77%, respectively.

The top three most important questions for cluster allocation were 1. “I still enjoy the things I used to enjoy”, 2 “Worrying thoughts go through my mind”, and 3. When I’m in pain it’s awful and I feel that it overwhelms me.”

## Discussion

Treatment failure in chronic pain patients is very common. Our aim was to identify subgroups of patients that are more or less likely to respond to certain interventions, so we can tailor subgroup-specific treatments to improve pain management. Based on the HADS, PCS and TSK questionnaires, we identified three chronic pain subgroups in a heterogeneous patient population (n=5,454) in two tertiary outpatient settings using hierarchical cluster analysis. Cluster 1 was characterized by high psychological burden, more tobacco smoking, lower educational levels, lower employment rates and more singles. Cluster 2 showed low psychological burden, more alcohol consumption, higher educational levels and higher employment rates. Cluster 3 showed intermediate features compared to the other clusters. Pain intensity and pain characteristics did not differ appreciably between the clusters in both cohorts. Regarding pain diagnosis, in cluster 1 widespread pain (UMCU cohort) and tendomyogenic pain (EMC cohort) were most prevalent. We hypothesize that patients with these diagnoses belong to a comparable diagnosis group with multifocal poorly defined chronic pain and that due to challenges in the classification of this pain syndrome this was either classified as widespread pain or tendomyogenic pain in the two centers. Complex regional pain syndrome (EMC cohort) was most prevalent in cluster 2. For all other diagnoses, there were no clinically relevant differences in prevalence among clusters. Importantly, treatment success was comparable between clusters 2 and 3, but was consistently and significantly lower in cluster 1. We hypothesize that patients identified as belonging to cluster 1 may need a different treatment approach, with suggestions provided below.

When comparing our findings with the current literature on cluster analyses of populations with chronic pain, the majority of studies also report differences observed in the psychological domain. Some of these studies found similar ‘extreme’ groups regarding psychological characteristics and pain experience, similar to our clusters 1 and 2, along with one or more intermediate group(s) like our cluster 3 [5,7–12,26]. The number of clusters differs between studies, varying from 2 to 9 clusters. Discrepancies in the number of clusters between different studies might be due to differences in the study populations or clustering variables used as input variables. Our identification of three clusters in a heterogenous chronic pain population (in the combined dataset and in both cohorts separately; see S2A and S2B Fig) is consistent with the findings of Gerdle et al. [12] and Gilam et al[26]. Gerdle et al. identified three groups based on variables of pain intensity, emotional distress, acceptance and life impacts. Their study revealed a group with overall ‘worst’ characteristics that included fewer subjects with a university education, similar to our cluster 1; a contrasting group with ‘best’ characteristics and with the highest proportion of subjects with university education, similar to our cluster 2, and an intermediate group similar to our cluster 3. Gilam et al. identified three groups based on three domains: physical, mental and social. In line with our clusters, the three groups showed graded severity in all of these domains and associated pain characteristics. Moreover, mental symptoms of anxiety, depression and anger were found to be the key determinants of subgroup assignment. Similarly, in our study, anxiety and depression belonged to the variables with the highest determinant value between all groups. Some studies added pain diagnoses in the clustering analysis. In contrast to our study, Bäckryd et al. [10] identified four subgroups and found an asymmetrical distribution of different diagnoses across groups, although these differences were small. Reviewing all these studies and including the observations in our own two large cohorts, we conclude that identification of three subgroups with a graded severity of psychological symptoms appears to be a robust finding.

Associated with these psychological symptoms are elements within the social domain, including educational level, employment status, life style factors and marital status. Unemployment, reducing socioeconomic status, may induce psychological stress [27,28]. Several studies have also shown that lower education levels and lower socioeconomic status correlate with higher pain prevalence and lower health status [29–31]. There are several hypothetical explanations for this phenomenon: People with less extensive education are more likely to be exposed to clinical, behavioral and environmental risk factors; they might have more physically demanding jobs and/or more unhealthy lifestyle behaviours; they might not have access to health care; and they might be more exposed to stress factors while also having poorer coping skills [32]. Health literacy possibly also plays a role in the underlying mechanism driving the interaction between low level of education and poor health [33]. A study on health literacy with n=131 chronic pain patients found that 54% had an inadequate health literacy and that this was associated with lower education level and lower monthly income [30]. In our study we also observed that cluster 1 was most associated with lower educational degree and least associated with university education and paid employment, in stark contrast to cluster 2. Our subgroups also differed regarding marital status, with more singles in cluster 1 compared to cluster 2. It has been previously suggested that social support by romantic partners might have an analgesic effect [31]. This study suggested that distress during pain exposure could be relieved by partner empathy, even solely with a partner’s physical presence, thereby reducing pain sensitivity and facilitating pain coping. Therefore, the lack of social support from a partner could perhaps be of influence on the pain experience of the patients in cluster 1. Furthermore, both psychological symptoms and high severity of pain can lead to a decreased HRQoL [34]. It is therefore not surprising that the identified groups showed such a pattern and that two groups (cluster 1 and cluster 2) emerged with highly contrasting characteristics.

In the current study the prevalence of <u>most</u> pain diagnoses did not differ between the three clusters, and the difference in pain intensity and characteristics between clusters were small and only significant in one of our two cohorts. This suggests that the actual initial inciting stimulus or painful condition may be of lesser importance to the chronic pain experience (duration, impact and severity) than psychosocial factors are. While the psychosocial factors may not be exclusive or specific for chronic pain, pain treatment outcomes were significantly different between cluster 1 and the other two clusters. This suggests that patients from cluster 1 present with a unique set of psychosocial factors that may need a different treatment approach. Possible changes in pain management could include pain education tailored to the educational level of the patient to improve understanding of their disease, lifestyle coaching including cessation of smoking (which is associated with higher pain intensity, pain interference and pain-related fear [35]), and support by social workers in finding a job and improving socioeconomic status to reduce stress that is associated with worse chronic pain. Naturally, patients with signs of a clinically significant anxiety or depressive disorder should be referred for psychiatric care, but we want to emphasize that this alone might not be enough, as the multidisciplinary treatment offered at the EMC (and also UMCU) included psychological and/or psychiatric referral when indicated.

There were three diagnoses: widespread pain, tendomyogenic pain and CRPS, which were not equally distributed across the clusters. Tendomyogenic pain and widespread pain are known to be associated with depression [36], which may explain the higher prevalence in cluster 1. CRPS is more often diagnosed in women which provides an explanation for the sex difference observed between both cohorts. In cluster 2, CRPS was overrepresented, for which no clear underlying cause could be identified. This overrepresentation warrant further investigation into possible underlying factors or covariates within this specific cluster (e.g. lifestyle factors, educational level influencing coping strategy).

The differences in the overall diagnosis distribution between the two centers seem mainly related to differences in research focus and healthcare expertise; the EMC is an expertise centre for CRPS, explaining the larger number of CRPS patients in their cohort.

This study has several strengths, including a large heterogenous chronic pain sample included at two multidisciplinary tertiary pain centres, with variables representing the different potential drivers of chronic pain according to the biopsychosocial model. However, this study also has several limitations that must be considered. First, approximately one third of patients had to be excluded from the hierarchical cluster analysis due to one or more missing values in the questionnaires. This large proportion of missing data might have biased the results, when patients not willing or not able to fill out all the questions are overrepresented in the excluded group. Second, the present results are based on a group of patients referred to a tertiary academic pain clinic, which tend to represent the most complex cases. Therefore, our findings may not generalize to other chronic pain populations and should be verified across different chronic pain patients and in different clinical settings. Third, due to the cross-sectional design of this study, causality cannot be determined. Lastly, the results are based on self-reported outcomes and could be biased by social desirability. People with higher educational levels may be more successful in manipulating their answers to questionnaires (such as the HADS, PCS and TCK) to reduce their psychological burden result and prevent a possible referral to the psychiatrist or psychologist when the patient is not motivated or open to such intervention.

Regarding clinical implications, the present study underlines the importance of acknowledging that the chronic pain population is not a homogenous group, and indicates that therapeutic interventions should be adjusted to individual patient characteristics rather than only to pain diagnoses. Subgroup assignment through psychometric questionnaires can potentially help support clinical decision making by clinicians: Using the knowledge of subgroup patterns, it can be decided which treatment options are most likely to be successful and therefore would be more preferable for the individual patient. It seems particularly important to identify patients that belong to cluster 1, as patients with this subset of characteristics are likely at risk of high-impact chronic pain, which is associated with the most suffering, most unfavourable health outcomes, increased medical costs and increased opioid use and dosage [37,38]. With our prediction model we can reliably predict cluster 1 allocation with an optimal number of questions of 15, leading to an accuracy for cluster 1 of 91% with a sensitivity of 96% and specificity of 86%. In future trials, the clinical relevance and treatment responses of subgroup-specific pain management approaches must be further evaluated.

## Conclusion

In conclusion, using hierarchical cluster analysis on two population cohorts, this study identified three chronic pain subgroups with different psychological and sociodemographic characteristics based on patient-reported measures. Remarkably, these groups were largely unrelated to specific pain diagnoses. This knowledge can be potentially useful for tailoring subgroup specific treatment plans to improve chronic pain management for individual patients. Using our prediction model including 15 questions only, we can reliably predict cluster allocation, especially to cluster 1, identifying patients who need a biopsychosocial approach with tailored pain education. It would be useful to validate these results in different pain populations and clinical settings, to determine whether these subgroups are widely clinically relevant, and to compare subgroup responses to different pain management strategies.

## Data Availability

All data produced in the present work are contained in the manuscript

## Acknowledgments

We want to thank Ilona van Gent-Bijen, George Groeneweg, and Max Brenninkmeijer for their support with the data extraction; Pain clinic secretaries for providing technical support to patients when filling out the digital questionnaires; Patients for their efforts filling in the questionnaires.

## Supporting information

**S1 Fig.**
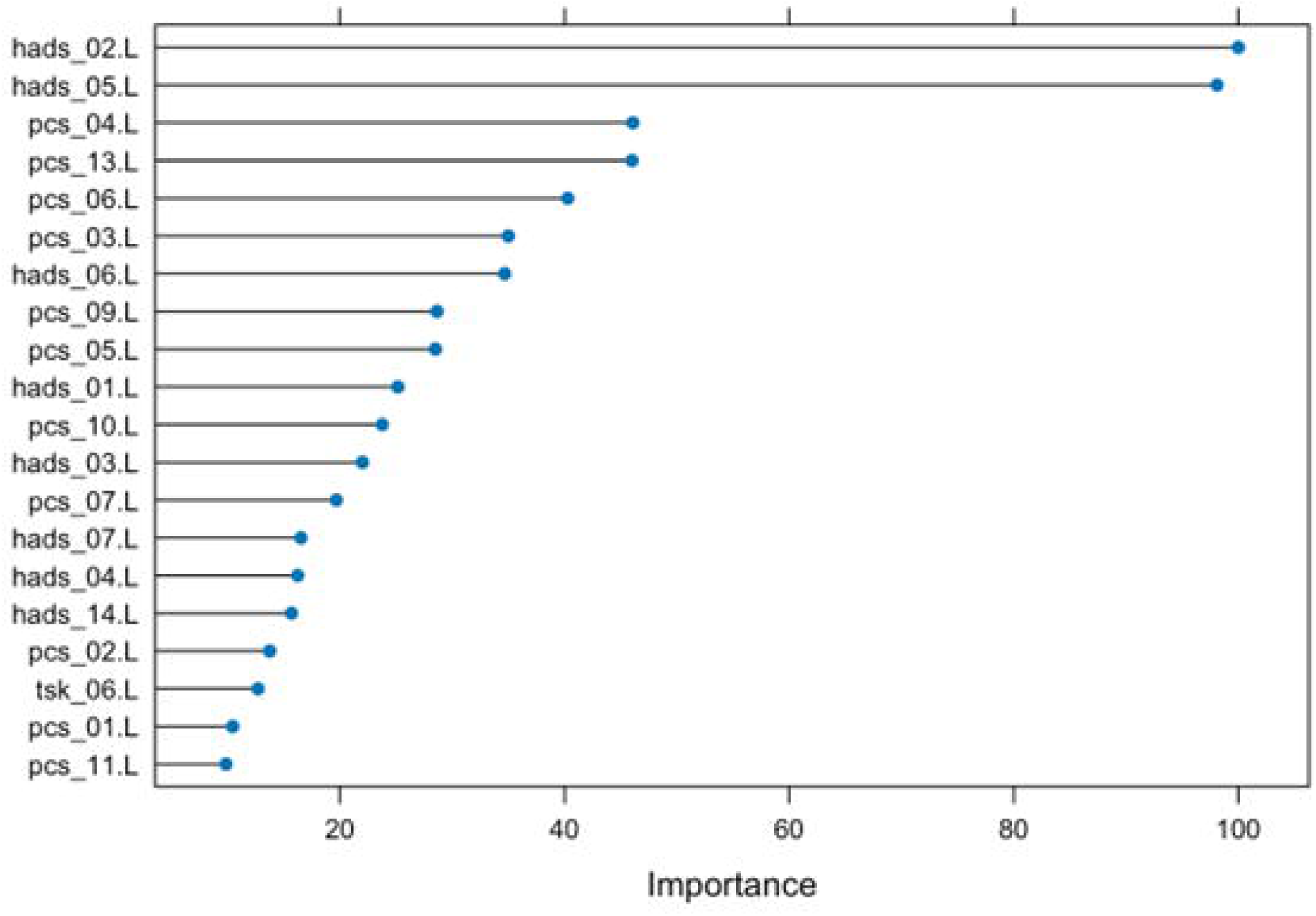
Top 20 questions. The 20 most important questions for accurate cluster prediction.

**S2A Fig.**
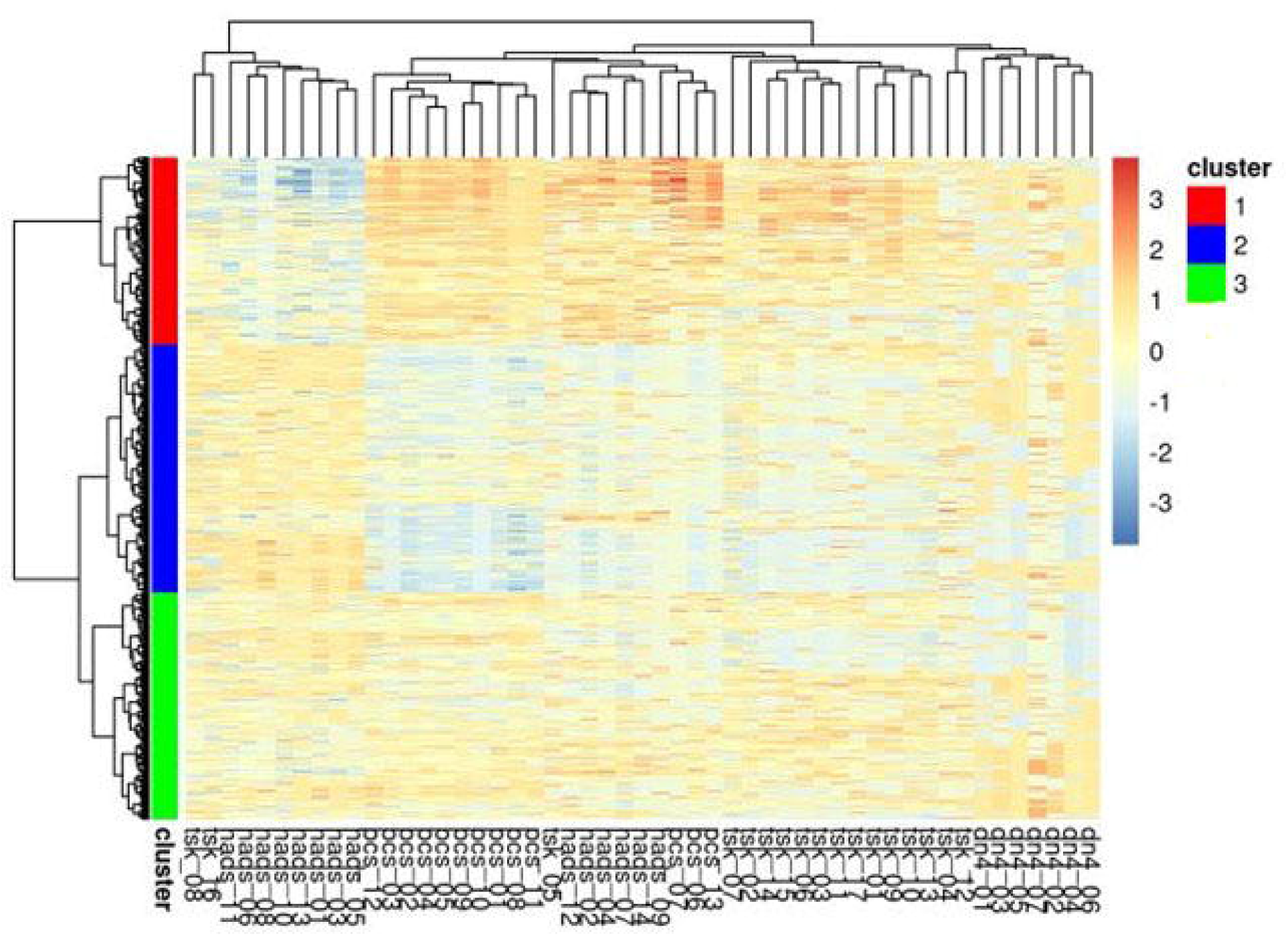
Hierarchical clustering dendogram of the development cohort University Medical Center Utrecht. Hierarchical cluster analysis showing three clusters: Cluster 1(red), Cluster 2 (blue) and Cluster 3 (green). X-axis representing the individual questions of three questionnaires (Hospital Anxiety and Depression Scale (HADS), Pain Catastrophizing Scale (PCS), and Tampa Scale of Kinesiophobia (TSK). Y-axis representing included patients.

**S2B Fig.**
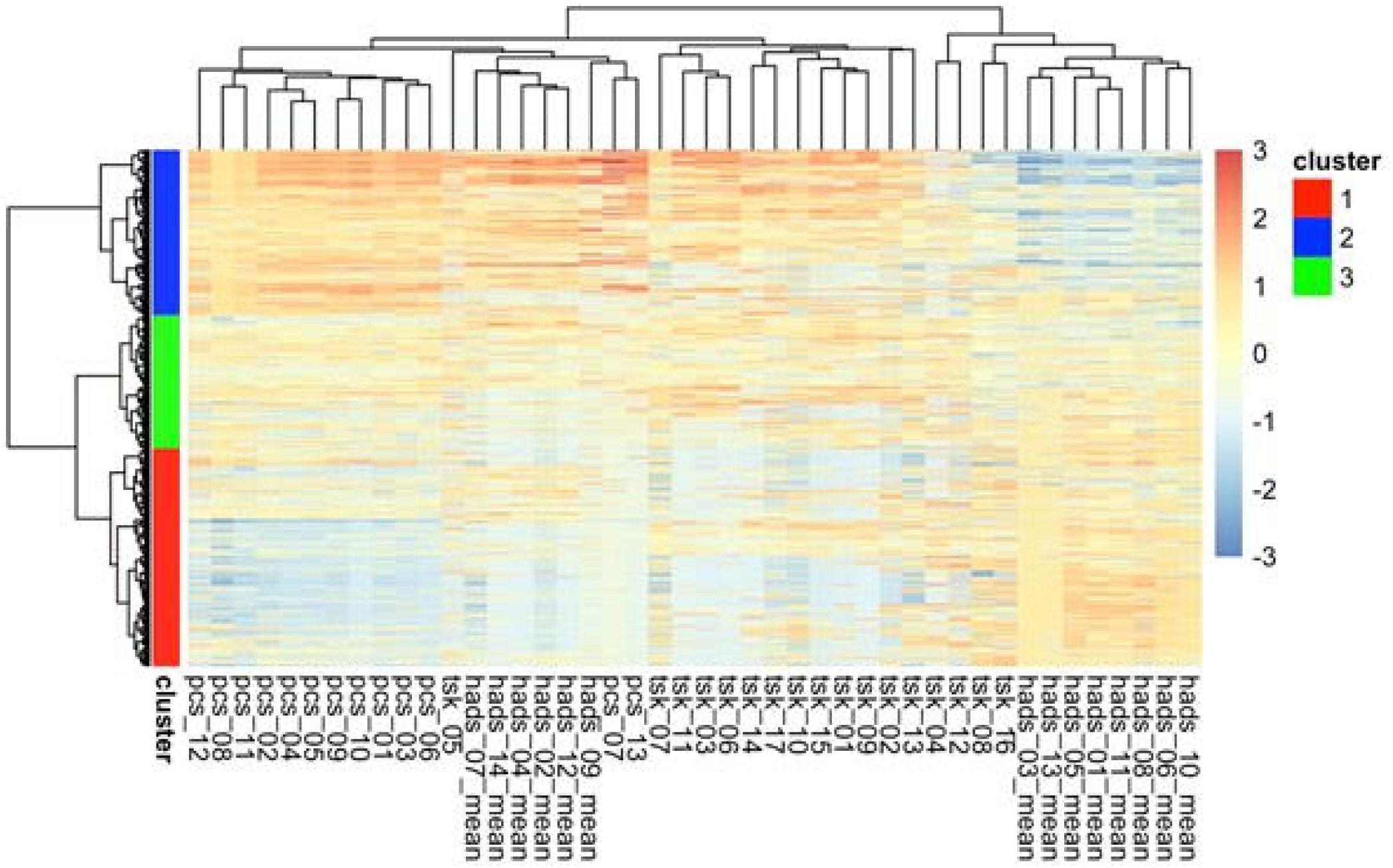
Hierarchical clustering dendogram of the validation cohort Erasmus MC. Hierarchical cluster analysis showing three clusters: Cluster 1(red), Cluster 2 (blue) and Cluster 3 (green). X-axis representing the individual questions of three questionnaires (Hospital Anxiety and Depression Scale (HADS), Pain Catastrophizing Scale (PCS), and Tampa Scale of Kinesiophobia (TSK). Y-axis representing included patients.

**S1 Table. Frequencies of the 10 most common diagnoses in each cluster (validation cohort Erasmus MC)**

Data expressed as percentage per cluster. Significance levels computed by Pearson-Chi square.

**S2 Table. Sociodemographics, pain intensity, duration, character, interference and health-related quality of life in the <u>validation cohort</u> (Erasmus Medical Center Rotterdam) and comparison among the clusters**

Data expressed as mean (95% confidence interval), median (interquartile range) or count (%). Statistics computed by one-Way ANOVA Test or Pearson Chi Square. P-values ≤ 0.01 are in bold. Effect sizes for categorical data are calculated with Eta-squared test or Cramer’s V. Effect sizes computed by Eta-squared test indicate a small effect 0.01 to 0.06, medium 0.06 to 0.14 and large effect ≥0.14. Effect sizes with Cramer’s V indicate a small effect 0.1 to 0.3, medium 0.3 to 0.5 and large effect >0.5. Significant differences with medium to large effect size are in bold.

Percentage of missing data: age 0%, gender 0%, BMI 29.9%, daily smoking 29.9%, highest educational level 0.2%, employment status 29.9%, marital status 0.2%, children 29.9%, pain NRS 29.9%, pain duration 1.3%, neuropathy (PD) 3.2%, BPI 28.8%, SF36 0%.

BMI: Body Mass Index; BPI: Brief Pain Inventory; HADS-A and HADS-D: Hospital Anxiety and Depression Scale; NRS: Numeric Rating Scale; PCS: Pain Catastrophizing Scale; PD: Pain Detect; SF36: Short Form-36; TSK: Tampa Scale for Kinesiophobia.

PCS subcategories represent the sum of following items ‘Rumination’ 8,9, 10 and 11; Magnification 6, 7 and 13; ‘Helplessness’ 1, 2, 3, 4, 5 and 12. RAND-36 represent the sum of the following items ‘General’ 1,11a,11b,11c,11d; ‘Physical’ 3a, 3b,3c,3d,3e,3f,3g,3h,3i,3j; ‘Mental’ 9b,9c,9d,9f,9h.

**S3A Table. Prediction characteristics for the validation cohort Erasmus MC using the 44 questions**

**S3B Table. Prediction characteristics for the validation cohort Erasmus MC using the 20 questions**

**S3C Table. Prediction characteristics for the validation cohort Erasmus MC using the 15 questions**

**S3D Table. Prediction characteristics for the validation cohort Erasmus MC using the 10 questions**

**S1 File. Methods section questionnaires**

