## Supplemental Methods for "Divergent treatment responses in chronic pain: Identifying subgroups of patients through cluster analysis"

**Supplementary file: Methods section**

*Hospital Anxiety and Depression Scale (HADS),*

This self-assessment questionnaire consists of a total of 14 questions with a subscale of seven items each for anxiety and depression (HADS-A and HADS-D), that are rated on 4-point numerical scale (0 to 3). Both subscale scores range from 0 to 21, with a higher score reflecting more psychological distress. The symptoms are considered clinically relevant for subscale scores above 8.

*Pain catastrophizing scale (PCS)*

Pain catastrophizing is measured with the PCS. This instrument assesses catastrophizing in three dimensions: magnification, rumination and helplessness. It consists of 13 pain-related statements on a 5 point scale with a total score ranging from 0-52. In this study the statements were rated from 1 to 5, therefore, the total score ranged from 13 to 65. A higher total score represents a higher level of pain catastrophizing.

*Tampa Scale of Kinesiophobia (TSK)*

Fear of movement and injury was assessed with the TSK questionnaire. It consists of 17 items that are rated on a 4-point scale, ranging from 1 (strongly disagree) to 4 (strongly agree). The total score ranges between 17 and 68, with higher scores indicating more pain-related fear.

*Health-related quality of life (HRQoL)*
During the study period clinical practice changed in the UMCU switching from the European Quality of Life instrument (EQ5D) to the Short Form-12 Health Survey (SF-12). Patients in the study therefore filled out either the EQ5D or the SF-12. In the EMC the Short Form-36 Health Survey (SF-36) was used.

SF-12 is a validated brief version of the SF-36 which assesses the general HRQoL. The SF12 consists of 12 questions and the SF36 of 36 questions that assess eight health domains for physical and mental health. Physical health domains include general health, physical functioning, role physical and body pain. Mental health domains include vitality, social functioning, role emotional and mental health. Each scale score is transformed into a 0 to 100 score, with higher values indicating a better physical and mental health.

EQ5D assesses patient’s perceived HRQoL through five dimensions: mobility, self-care, usual activities, pain/discomfort and anxiety/depression, with three optional answers for each item (‘no problems’, ‘some problems’, ‘major problems’). Using these dimensions, a country-specific index value is calculated (EQ5D index). A global self-estimate of health is also assessed on a thermometer-like scale ranging from 0-100 (EQ5D VAS), with high scores indicating a good health and low scores bad health.
