## Supplemental Tables for "Divergent treatment responses in chronic pain: Identifying subgroups of patients through cluster analysis"

**Supplementary tables**

**Table 1** Frequencies of the 10 most common diagnoses in each cluster (validation cohort Erasmus MC)

|  | Total | Cluster 1 | Cluster 2 | Cluster 3 | Χ^2^ (df) | p-value |
| --- | --- | --- | --- | --- | --- | --- |
| Top 10 Diagnoses (%) |  |  |  |  | 164 (56) | <0.001 |
| 1. Other neuropathic pain | 17.5% | 17.4% | 16.4% | 18.3% |  |  |
| 2. Radicular syndrome | 17.0% | 14.3% | 17.8% | 17.1% |  |  |
| 3. Tendomyogenic pain | 12.1% | 19.4% | 8.8% | 12.4% |  |  |
| 4. Complex regional pain syndrome (CRPS) | 11.4% | 5.1% | 15.7% | 10.1% |  |  |
| 5. Mechanical spine related pain | 10.2% | 9.8% | 11.1% | 9.8% |  |  |
| 6. Joint pain | 6.2% | 5.3% | 5.3% | 7.1% |  |  |
| 7. Abdominal pain | 4.8% | 3.0% | 4.7% | 5.3% |  |  |
| 8. Central neuropathic pain | 4.2% | 5.8% | 3.3% | 4.3% |  |  |
| 9. Thoracal pain | 3.9% | 3.2% | 3.9% | 4.0% |  |  |
| 10. Orofacial pain | 2.4% | 3.2% | 2.6% | 2.0% |  |  |

Data expressed as percentage per cluster. Significance levels computed by Pearson-Chi square.

**Table 2.** Sociodemographics, pain intensity, duration, character, interference and health-related quality of life in the validation cohort (Erasmus Medical Center Rotterdam) and comparison among the clusters

| **Variables** | | | **Total sample**  **(n=3855)** | **Cluster 1**  **(n=1235, 32.0%)** | **Cluster 2**  **(n=1625, 42.2%)** | **Cluster 3**  **(n=995, 25.8%)** | ***χ ^2^ (df)*** | ***p-value*** | ***Effect size*** |
| --- | --- | --- | --- | --- | --- | --- | --- | --- | --- |
| Age (y) | | | 49.9 (49.4-50.4) | 49.2 (48.4-50.1) | 49.4 (48.7-50.2) | 51.4 (50.4-52.4) |  | 0.002 | 0.003 |
| Gender (female) | | | 2478 (64.2%) | 57.0% | 70.3% | 63.2% | 54.9 (2) | <0.001 | 0.119 |
| BMI (kg/m^2^) | | | 24.2 (23.2-25.1) | 23.6 (23.2-24.0) | 23.5 (23.0-24.0) | 26.0 (22.5-29.5) |  | 0.085 | 0.002 |
| Daily smoking (yes) | | | 638 (23.6%) | 31.4% | 18.6% | 21.9% | 46.2 (2) | <0.001 | 0.131 |
| Highest educational level  -Primary education  -Secondary education  -Vocational education  -University  -Education not specified | | | 318 (8.3%)  825 (21.4%)  1727 (44.9%)  897 (23.3%)  80 (2.1%) | 14.1%  22.5%  43.5%  16.2%  3.7% | 4.6%  20.6%  45.8%  28.1%  0.9% | 6.9%  21.5%  45.1%  24.4%  2.0% | 151.3 (8) | <0.001 | 0.140 |
| Employment status  -Student  -Retired  -Unable to work  -Volunteer/unpaid employment  -Employed  -Unemployed  -Other | | | 114 (4.2%)  468 (17.3%)  527 (19.5%)  256 (9.5%)  1003 (37.1%)  284 (10.5%)  50 (1.8%) | 4.1%  17.0%  21.9%  8.1%  29.5%  17.4%  2.1% | 5.0%  14.9%  19.9%  10.8%  42.2%  5.3%  1.9% | 3.1%  21.5%  15.9%  9.1%  38.5%  10.3%  1.5% | 121.3 (14) | <0.001 | 0.150 |
| Marital status  -Single  -Cohabitation/marriage/ widowed  -Other | | | 956 (24.9%)  2501 (65%)  390 (10.1%) | 29.6%  59.7%  10.7% | 21.0%  68.3%  10.6% | 25.2%  66.2%  8.6% | 32.3 (4) | <0.001 | 0.065 |
| Children (yes) | | | 1842 (68.2%) | 70.4% | 66.2% | 68.5% | 4.1 (2) | 0.127 | 0.039 |
| Average pain (NRS) | | | 8(2) | 7(2) | 8(1) | 8(1) |  | <0.001 | 0.085 |
| Min. Pain (NRS) | | | 5(4) | 5(3) | 6(2) | 5(3) |  | <0.001 | 0.088 |
| Max. Pain (NRS) | | | 9(1) | 9(2) | 10(1) | 9(1) |  | <0.001 | 0.068 |
| Pain duration (%) ≥ 1 year | | | 86.4% | 86.3% | 87.4% | 85.5% | 1.8(2) | 0.407 | 0.022 |
| Neuropathy (PD) >90% Certainty (%) | | | 39.9% | 31.9% | 50.8% | 39.5% | 116(4) | <0.001 | 0.125 |
| Brief Pain Inventory | | |  |  |  |  |  |  |  |
| General Activity | | | 7(2) | 7(3) | 8(2) | 7(2) |  | <0.001 | 0.072 |
| Mood | | | 7(3) | 6(4) | 8(2) | 7(3) |  | <0.001 | 0.150 |
| Walking ability | | | 7(5) | 6(5) | 8(3) | 7(4) |  | <0.001 | 0.046 |
| Normal Work | | | 8(3) | 7(4) | 9(2) | 8(3) |  | <0.001 | 0.053 |
| Relations with other people | | | 6(5) | 4(6) | 7(3) | 6(5) |  | <0.001 | 0.136 |
| Sleep | | | 7(4) | 7(4) | 8(3) | 7(4) |  | <0.001 | 0.047 |
| Enjoyment of life | | | 7(5) | 5(4) | 9(3) | 7(3) |  | <0.001 | 0.172 |
| Quality of life (SF36) | | |  |  |  |  |  |  |  |
| Physical | | | 31.9 (31.6-32.1) | 30.5 (30.2-30.9) | 33.2 (32.7-33.6) | 31.4 (31.0-31.9) |  | <0.001 | 0.021 |
| Mental | | | 45.1 (44.8-45.4) | 38.2 (37.6-38.7) | 49.9 (49.5-50.3) | 45.9 (45.3-46.5) |  | <0.001 | 0.219 |

Table 1. Data expressed as mean (95% confidence interval), median (interquartile range) or count (%). Statistics computed by one-Way ANOVA Test or Pearson Chi Square. P-values ≤ 0.01 are in bold. Effect sizes for categorical data are calculated with Eta-squared test or Cramer’s V. Effect sizes computed by Eta-squared test indicate a small effect 0.01 to 0.06, medium 0.06 to 0.14 and large effect ≥0.14. Effect sizes with Cramer’s V indicate a small effect 0.1 to 0.3, medium 0.3 to 0.5 and large effect >0.5. Significant differences with medium to large effect size are in bold.

Percentage of missing data: age 0%, gender 0%, BMI 29.9%, daily smoking 29.9%, highest educational level 0.2%, employment status 29.9%, marital status 0.2%, children 29.9%, pain NRS 29.9%, pain duration 1.3%, neuropathy (PD) 3.2%, BPI 28.8%, SF36 0%.

BMI: Body Mass Index; BPI: Brief Pain Inventory; HADS-A and HADS-D: Hospital Anxiety and Depression Scale; NRS: Numeric Rating Scale; PCS: Pain Catastrophizing Scale; PD: Pain Detect; SF36: Short Form-36; TSK: Tampa Scale for Kinesiophobia.

PCS subcategories represent the sum of following items ‘Rumination’ 8,9, 10 and 11; Magnification 6, 7 and 13; ‘Helplessness’ 1, 2, 3, 4, 5 and 12. RAND-36 represent the sum of the following items ‘General’ 1,11a ,11b ,11c ,11d; ‘Physical’ 3a, 3b,3c,3d,3e,3f,3g,3h,3i,3j; ‘Mental’ 9b,9c,9d,9f,9h.

Table 3A. Prediction characteristics for the EMC cohort using the 44 questions

|  | Cluster 1 | Cluster 2 | Cluster 3 |
| --- | --- | --- | --- |
| Sensitivity (%) | 97.4 | 76.2 | 61.4 |
| Specificity (%) | 86.0 | 96.4 | 79.4 |
| Positive Predictive Value (%) | 40.6 | 96.1 | 63.8 |
| Negative Predictive Value (%) | 99.7 | 77.7 | 77.6 |

Table 3B. Prediction characteristics for the EMC cohort using the 20 questions

|  | Cluster 1 | Cluster 2 | Cluster 3 |
| --- | --- | --- | --- |
| Sensitivity (%) | 95.9 | 74.4 | 58.6 |
| Specificity (%) | 85.3 | 95.6 | 77.5 |
| Positive Predictive Value (%) | 37.2 | 95.3 | 60.5 |
| Negative Predictive Value (%) | 99.6 | 75.5 | 76.2 |

Table 3C. Prediction characteristics for the EMC cohort using the 15 questions

|  | Cluster 1 | Cluster 2 | Cluster 3 |
| --- | --- | --- | --- |
| Sensitivity (%) | 95.9 | 73.7 | 58.3 |
| Specificity (%) | 85.7 | 94.7 | 77.1 |
| Positive Predictive Value (%) | 39.5 | 94.3 | 59.4 |
| Negative Predictive Value (%) | 99.5 | 75.0 | 76.2 |

Table 3D. Prediction characteristics for the EMC cohort using the 10 questions

|  | Cluster 1 | Cluster 2 | Cluster 3 |
| --- | --- | --- | --- |
| Sensitivity (%) | 91.7 | 73.3 | 58.1 |
| Specificity (%) | 86.4 | 93.1 | 76.7 |
| Positive Predictive Value (%) | 43.1 | 92.5 | 58.4 |
| Negative Predictive Value (%) | 98.9 | 74.8 | 76.5 |
